# From Bedside to Bench: *Drosophila* Models of Baker-Gordon Syndrome (BAGOS)

**DOI:** 10.64898/2026.04.04.26350071

**Authors:** Carlos E. Rivera, Jenny Park, Brandon L. Holder, Lily Mattingly, Opal Ao, Christopher L. Anderson, Lucia T. Carney, Daniel J. Davis, Benjamin T. Black, Stephane Dissel, Paul R. Carney, Bing Zhang

## Abstract

*De novo SYT1* mutations cause Baker–Gordon syndrome (BAGOS), yet the pathogenic mechanisms are not well understood. We identified a child carrying a newly described *SYT1* variant, D310N, which produced a consistently more severe clinical phenotype compared to a previously reported D366E variant. To investigate the biological basis of these differences, we generated *Drosophila* models harboring each variant. Heterozygous D310N flies displayed substantially greater locomotor impairment, higher incidences of seizure-like activity, more pronounced deficits in learning and memory, and synaptic dysfunction than flies expressing D366E. Importantly, we report the identification of a mid-larval developmental window during which variant expression induces life-long locomotor abnormalities. Yet, mutant SYT1 expressed in adult stage does not have an immediate effect on climbing, arguing that BAGOS is likely caused by developmentally disrupted networks rather than synaptic transmission alone. Finally, we show behavioral abnormalities specific to SYT1 variant expression in subset neurons. Together, findings from this integrated human-fly analysis recapitulate core clinical manifestations, uncover variant-specific disruptions in SV recycling, developmental timing, and circuit-level contributions, and advance understanding of *SYT1*-associated neurodevelopmental disorders.

## Introduction

Neurotransmission at chemical synapses is essential for brain function. When an action potential arrives at the presynaptic terminal, calcium (Ca²⁺) rapidly enters through voltage-gated channels and triggers synaptic vesicle fusion and transmitter release. This process depends critically on Synaptotagmin-1 (SYT1), a vesicle-associated Ca²⁺ sensor that enables rapid and synchronous exocytosis by coupling Ca²⁺ binding to membrane fusion. The indispensable role of SYT1 in Ca²⁺-evoked neurotransmitter release has been firmly established through biochemical, electrophysiological, and genetic studies (*1–7*).

Given its essential function as the primary Ca²⁺ sensor for fast synaptic vesicle release, it is not surprising that SYT1 plays a critical role in human health. *De novo* mutations in *SYT1* have been identified as the cause of Baker-Gordon Syndrome (BAGOS), a rare neurodevelopmental disorder (*8–11*). Individuals with BAGOS exhibit a broad range of developmental challenges, including impairments in locomotion, sleep, digestion, vision, and cognition. To date, at least 18 pathogenic *SYT1* variants have been reported, all arising *de novo* and clustering within the C2A or C2B domains. These mutations manifest clinically in the heterozygous state, consistent with dominant-negative effects on SYT1 function. Emerging functional studies, largely conducted in cultured neurons (*9, 10, 12–15*), consistently reveal synaptic defects aligned with the established roles of SYT1 in both SV exocytosis and endocytosis.

At present, no viable animal model of BAGOS exists, limiting opportunities to investigate the disorder at developmental, behavioral, and neural-circuit levels. The pathogenic mechanisms linking *SYT1* mutations to disease are not well understood, and no therapeutic interventions have been developed. *Drosophila* provides a powerful platform for dissecting synaptic physiology and behavior due to its simplified but highly conserved nervous system and genetic architecture. In this study, we integrated clinical observations from two children with *Drosophila* models carrying their corresponding *SYT1* variants to elucidate physiological and behavioral mechanisms. A newly identified variant, D310N, was compared with a previously reported pathogenic variant, D366E. The fly models recapitulated key patient features, including locomotor impairment, seizure-like activity, ataxia, and memory deficits. Studies of our fly models also revealed a critical developmental window during which variant expression drives persistent behavioral abnormalities and the absence of phenotype when mutant *SYT1* is expressed only in mature animals. These findings imply a defect beyond neurotransmission *per se*; rather it argues for developmental abnormalities of neural networks.

## Materials and Methods

### Identification of Syt1 Patients

Human participants with *SYT1* missense variants were evaluated using the standardized assessment framework of the Baker-Gordon Syndrome (BAGOS) natural history study (ClinicalTrials.gov Identifier: NCT06399952). The protocol employs a multimodal battery to characterize developmental, behavioral, motor, and neurophysiologic features of *SYT1*-associated neurodevelopmental disorder, modeled after approaches used in recent natural-history studies of rare neurodevelopmental syndromes (*9, 16*). All clinical assessments were administered by trained clinicians or certified psychometric evaluators using age-normed, standardized procedures.

### Clinical evaluations of *SYT1* patients

All evaluations were conducted under an Institutional Review Board–approved protocol, with written informed consent obtained from each participant’s legal guardian in accordance with institutional and federal guidelines. Individuals with SYT1 missense variants were evaluated using research-reliable, age-normed psychometric instruments, with all data reviewed and interpreted by an experienced clinician. Assessments were administered by trained clinicians or certified psychometric evaluators following standardized procedures.

Developmental and cognitive functioning were assessed using the Mullen Scales of Early Learning (MSEL) (*17*). Adaptive functioning was evaluated via caregiver interview using the Adaptive Behavior Assessment System, Third Edition (ABAS-3) (*18*). Motor function was characterized through focused neurologic examination and functional skill assessment (*19*). Behavioral and neuropsychiatric features were assessed using standardized caregiver-reported measures, including the Child Behavior Checklist (CBCL) (*20*), Social Responsiveness Scale-second edition (SRS-2) (*21*), Repetitive Behavior Scale-Revised (RBS-R) (*22*), Aberrant Behavior Checklist (ABC) (*23*), and Vanderbilt Assessment Scale (*24*), as age-appropriate.

The ABAS-3 was selected as the primary adaptive behavior measure to allow consistent assessment across the studied age range and to facilitate comparison of caregiver-reported functional skill domains emphasized in prior studies of SYT1-associated and related neurodevelopmental disorders.

Sleep patterns and paroxysmal episodes were monitored through caregiver diaries documenting sleep duration, nighttime awakenings, and transient motor or behavioral events. Home video recordings, when available, were reviewed to contextualize episodes and differentiate behavioral events from possible seizure-like activity, consistent with natural-history methodologies in neurogenetic epileptic encephalopathies (*25, 26*).

### Generation of CRISPR–Cas9–edited *Drosophila syt1* mutant lines

To model patient-specific mutations in the endogenous *Drosophila syt1* locus, orthologous amino-acid substitutions were introduced using CRISPR-Cas9 mediated genome modification.

Two variants were generated: *syt1*-D362N (named *dsyt1^D362N^*), corresponding to the human *SYT1*-D310N mutation, and *syt1*-D418E (*dsyt1^D418E^*), corresponding to the human *SYT1*-D366E mutation.

Single-guide RNAs (sgRNAs) were designed to target sequences adjacent to the desired codons, and donor templates carrying the respective nucleotide substitutions were synthesized. Embryo injections were performed by a commercial service (Rainbow Transgenics), using a Cas9-expressing strain for germline editing. Putative edited founders (G₀) were crossed individually to balancer stocks, and G₁ progeny were screened by PCR amplification of the targeted *syt1* region followed by sequence analysis to identify precise HDR-derived alleles. For each mutation, independent lines carrying the correct nucleotide substitution and no additional coding changes were isolated. Mutant chromosomes were balanced, and stable stocks were expanded and maintained under standard conditions. Final sequences of both *syt1* variants were verified by Sanger sequencing of genomic PCR products spanning the edited region.

### Generation of *Drosophila* lines expressing human *SYT1* mutant transgenes

Patient-specific human *SYT1* variants were engineered by site-directed mutagenesis of the full-length human *SYT1* cDNA. Individual point mutations were introduced to generate the following alleles, D310N and D366E. Each mutant cDNA was cloned into the *phiC31*-integrase compatible vector pUASTB, with *attB*, which enables targeted genomic insertion onto third chromosome marked with *attP* at location 86Fb.

Constructs were verified by Sanger sequencing and submitted to Rainbow Transgenics (Camarillo, CA) for microinjection. Transgenes were integrated into the standard *attP* landing site using the *phiC31* system. Resulting transformants were balanced using standard genetic strategies, and stable stocks were established. For each line, genomic DNA was isolated from adult flies, and site-specific insertion and sequence fidelity of the integrated transgene were confirmed by PCR amplification followed by sequencing across the junction and coding regions.

### Fly genetics and husbandry

Additional fly lines were obtained from the Bloomington *Drosophila* Stock Center, including C155-Gal4 (BL#458), Elav-GSW-Gal4 (BL#43642), GAD1-Gal4 (BL#51630), vGlut-Gal4 (BL#24635), Ddc-Gal4 (BL#7010), Chat-Gal4 (BL#6793), and Ok6-Gal4 (BL#64199). Unless otherwise noted, flies and genetic crosses were maintained in incubators at 25°C on a 12:12 light–dark cycle and reared on standard cornmeal medium in polypropylene vials. The fly food contained dextrose (8% w/v), cornmeal (3.8% w/v), inactive yeast (2% w/v), agar (0.5% w/v), and tegosept (0.16% w/v).

Major fly genetic crosses are briefly described here. *dsyt1^D362N^*and *dsyt1^D418E^* were placed on compound balancers as (*dsyt1^D362N^*; +)/CyO-Tb or (*dsyt1^D418E^*; +)/CyO-Tb. Homozygotes of *dsyt1^D362N^*were not possible because they were embryonic lethal. Homozygotes of *dsyt1^D418E^*were collected as non-Tb larvae or non-CyO and non-Tb adult flies. To facilitate the collection of homozygotes, (*dsyt1^D418E^*; +)/CyO-Tb flies were kept at 18°C per protocol established by the Reist lab (*27*). Heterozygotes were generated by crossing (*dsyt1^D362N^*; +)/CyO-Tb or (*dsyt1^D418E^*; +)/CyO-Tb to CS flies.

Three to five independent insertion lines for the transgenic UAS-*SYT1* variants or CRISPR mutants were recovered and underwent a pilot screen for climbing defects. They showed similar defects and thereafter one line was selected for further analysis. To express *SYT1* variants in different neurons, the Gal4 lines were crossed to UAS-*SYT1* variants and the F1 were collected and examined for behavioral, functional, and morphological studies. UAS-*SYT1* variants/+ and Gal4/+ were used as control groups.

### Climbing assay

The climbing assay was performed by adapting the method described previously (*28–30*). Adult flies were briefly anesthetized under CO₂ within 0–6 hours after eclosion and separated by sex into vials containing approximately 10–20 flies each. Flies were aged for 5 and/or 10 days and transferred to fresh food vials regularly. For behavioral testing, flies were placed into an empty 100-ml graduated cylinder, which was capped with a cotton plug to prevent escape. Cylinders were secured to a frame, lifted from the bench, and tapped firmly several times to bring all flies to the bottom. Climbing behavior was video recorded using either a Samsung Galaxy S22 Ultra or an iPhone 13 camera. For analysis, the distance each fly traveled during the first 10 seconds was measured, and these values were used to calculate average climbing speeds.

### Bang-sensitive assay

Experiments were performed similarly as described (*31, 32*). Briefly, 7-day old flies were placed in empty plastic tubes and vortexed at high speed for 20 seconds. Immediately after vortexing, flies were video recorded for up to 30 seconds.

### Other locomotive behaviors

Some flies also displayed seizure-like activities during the climbing test and bang-sensitive assay. These flies buzzed their wings uncontrollably or bounced around inside the vial, and at times fell on their backs and struggled to upright themselves. Further, ataxia-like behavior defined as a sluggish/staggering motion in their walking gate was also observed.

### Larval crawling

Wandering-stage larvae were transferred onto a large Petri dish coated with a thin layer of 1% agarose (*33*). The dish was placed in a recording chamber illuminated from below by an LED light source. Three larvae were positioned near the center of the dish, approximately one inch apart, and allowed to crawl freely. Crawling velocity was quantified using the wrMTrck plugin running in Fiji. Videos were captured with a Point Grey CMOS camera equipped with a Fujinon DF6HA-1B lens.

### GeneSwitch experiments

The GeneSwitch experiment was performed following the methods described previously (*34, 35*). RU486 was purchased from Sigma-Aldrich, Inc (St Louis, MO) and dissolved in ethanol. To make the RU486 food, RU486 was added to the normal fly food at a final concentration of 20 µM. The vehicle food (control) contained the same amount of ethanol equivalent to that was used to make the 20 µM RU486 solution.

### Electrophysiology

Third-instar larvae were used for all electrophysiological experiments using neuromuscular junction preparation (*36*) and established methods in the Zhang lab (*37–39*). HL-3 solution (*40*) containing 0.8 mM Ca²⁺ was used for routine recordings, whereas 1.5 mM Ca²⁺ was used for depletion assays. For dissections, larvae were pinned at the head and tail and gently stretched. A midline incision was made, and four additional pins were used to secure the body wall. Internal organs were removed, leaving the musculature and nervous system intact. Borosilicate microelectrodes (Precision Instruments 1B100F-4) were pulled on a Sutter P-2000 puller and had tip resistance of approximately 50 mΩ. Recording electrodes were inserted into muscle 6 or 7 of abdominal segments A2 or A3.

For evoked excitatory junctional potential (EJP) and miniature event recordings, segmental nerves were severed rostrally near the tip of the ventral nerve cord using Vannas ROBOX scissors. A suction electrode was fabricated by breaking and fire-polishing the tip of a pulled pipette to create an opening large enough to enclose a single nerve. The nerve was gently aspirated into the electrode using a 1-ml syringe. Stimulation was delivered every 5 seconds with a Master-8 (A.M.P.I), and signals were acquired with an AxoClamp 2B amplifier and pClamp software. Muscle input resistance was measured routinely, and preparations with values < 8 mΩ were excluded. For depletion experiments, nerves were stimulated at 10 Hz for 5 minutes. Data shown as ±SEM.

### Immunohistochemistry

The general procedure of immunocytochemistry was adapted as described earlier (*41–43*). Third-instar larvae were dissected in ice-cold, zero-Ca²⁺ HL-3 on a small dish lined with 10% Sylgard. Immediately after dissection, tissues were transferred to Bouin’s fixative for 30 minutes, then washed in PBS for 10 minutes in 1.5-ml Eppendorf tubes. Samples were subsequently washed three times in PBST for 15 minutes each before being incubated overnight at 4 °C in primary antibodies. Mouse anti-DLG (1:1000; DSHB), rabbit anti-dSyt1 (1:500), and rabbit anti–human SYT1 (1:8000; Fisher Scientific PA5-29922) were used.

After an overnight incubation with primary antibodies, tissues were washed three times in PBST for 15 minutes each and then incubated with secondary antibodies diluted 1:1000. Alexa Fluor 488 donkey anti-rabbit and Alexa Fluor 594 goat anti-mouse antibodies were purchased from Fisher Scientific. Samples were mounted on microscope slides and initially examined using an Olympus BX61 motorized microscope equipped with a BX-DSU unit. Final images were acquired on a Leica TCS SP8 system with digital light-sheet and diode laser illumination.

### Learning and memory tests

The courtship learning and memory assay was performed according to the methods described (*44, 45*). Naïve males were collected at eclosion, isolated into individual glass tubes, and aged for 4–5 days. For short-term memory (STM) assays, a single male was placed into a mating chamber with a mated female for a 1-hour training session. Males were then returned to their glass tubes for a 30-minute rest period before being transferred to courtship testing chambers with a new mated female and recorded for 10 minutes.

For long-term memory (LTM) assays, single males were paired with a mated female for a 6-hour training session. After training, males were returned to glass tubes for approximately 18 hours.

Roughly 24 hours after the onset of training, males were transferred to courtship testing chambers with a new mated female and recorded for 10 minutes.

Videos were analyzed using FlyTracker software (*46, 47*), and behavioral outputs were classified using JAABA (*48*) to compute courtship indices. Courtship index (CI) was calculated as CI = (time engaged in courtship behavior/ total test duration) X 100. Suppression index (SI) was calculated as SI = 100 X (1-CI_trained/CI untrained_average).

### Molecular dynamics simulations

To assess variant-specific structural effects on SYT1 function, molecular dynamics (MD) simulations were performed on the SYT1 C2B domain containing either the wild-type sequence or the patient-derived variants D310N and D366E. High-resolution structural models were generated with Ca²⁺ ions placed in canonical binding sites, solvated in explicit water with physiological ionic strength, and simulated under standard NPT conditions. Multiple independent trajectories were generated for each genotype to evaluate structural stability, local flexibility, and Ca²⁺-binding loop dynamics. Analyses focused on backbone deviation, residue-level flexibility, Ca²⁺ coordination geometry, and dominant modes of motion involving the Ca²⁺-binding loops. Full simulation protocols and extended analyses are provided in the Supplementary Materials.

## Data analysis

All climbing and electrophysiological data were analyzed using RStudio. Normality was assessed using the D’Agostino test. For nonparametric analyses, Kruskal–Wallis tests were performed followed by Dunn’s post hoc comparisons. For parametric analyses, ANOVA was used with Tukey post hoc tests. Learning and memory data were evaluated using the D’Agostino–Pearson test for normality; datasets that passed were analyzed with parametric tests, whereas those that failed were analyzed with nonparametric methods. Courtship index values were compared using Mann–Whitney U tests, and suppression index values were evaluated using one-sample Wilcoxon tests. Climbing, crawling, and physiology data with the UAS-SYT1 variants data were considered significantly different if the experimental line was significantly different than both parental controls (Gal4/+ and UAS/+).

## Results

### Clinical characterization of two individuals with BAGOS (SYT1) missense variants

Both individuals with *SYT1* missense variants demonstrated severe global neurodevelopmental impairment across developmental, adaptive, motor, and behavioral domains (**Tables 1 and 2**).

**Table 1.** Developmental, adaptive, and behavioral assessment results for a female in the 6–10 year age range with the SYT1 p.Asp366Glu (D366E) variant.

| DEVELOPMENTAL FUNCTIONING: Mullen Scales of Early Learning |  |  |  |
| --- | --- | --- | --- |
|  | Standard Score |  | Range |
| Early Learning Composite | <49 |  | Exceptionally Low |
| Scale | T-Score | Age Equivalent | Score Label |
| Visual Reception | <20 | 16 months | Exceptionally Low |
| Fine Motor | <20 | 23 months | Exceptionally Low |
| Receptive Language | <20 | 13 months | Exceptionally Low |
| Expressive Language | <20 | 15 months | Exceptionally Low |
| ADAPTIVE FUNCTIONING: ABAS-3 – Parent Form |  |  |  |
| Composite | Standard Score |  | Range |
| General Adaptive | 50 |  | Exceptionally Low |
| Conceptual | 51 |  | Exceptionally Low |
| Social | 55 |  | Exceptionally Low |
| Practical | 56 |  | Exceptionally Low |
| Skill Area | Scaled Score |  | Range |
| Communication | 01 |  | Exceptionally Low |
| Community Use | 01 |  | Exceptionally Low |
| Functional Academics | 01 |  | Exceptionally Low |
| Home Living | 04 |  | Below Average |
| Health and Safety | 02 |  | Exceptionally Low |
| Leisure | 02 |  | Exceptionally Low |
| Self-Care | 01 |  | Exceptionally Low |
| Self-Direction | 01 |  | Exceptionally Low |
| Social | 01 |  | Exceptionally Low |
| Motor | 05 |  | Below Average |
| BEHAVIORAL AND EMOTIONAL FUNCTIONING: CBCL, 1 ½ to 5 years of age |  |  |  |
| Scale | T-Score |  | Range |
| Internalizing | 70 |  | Clinically Significant |
| Emotionally Reactive | 70 |  | Clinically Significant |
| Anxious/Depressed | 63 |  | Within Normal Limits |
| Somatic Complaints | 58 |  | Within Normal Limits |
| Withdrawn | 73 |  | Clinically Significant |
| Externalizing | 74 |  | Clinically Significant |
| Attention Problems | 77 |  | Clinically Significant |
| Aggressive Behavior | 70 |  | Clinically Significant |
| DSM-5 Problems |  |  |  |
| Depressive | 70 |  | Clinically Significant |
| Anxiety | 75 |  | Clinically Significant |
| Autism Spectrum | 76 |  | Clinically Significant |
| ADHD | 76 |  | Clinically Significant |
| Oppositional Defiant | 67 |  | At Risk |
| Sleep Problems | 53 |  | Within Normal Limits |
| Total | 76 |  | Clinically Significant |
| Social Responsiveness Scale (SRS-2) |  |  |  |
| Scale | Raw |  | T-Score |
| SRS total raw score | 114 |  | 78 (severe range) |
| Social Awareness | 16 |  | 74 |
| Social Cognition | 20 |  | 70 |
| Social Communication | 37 |  | 75 |
| Social Motivation | 15 |  | 66 |
| Restrictive and Repetitive Behaviors (RRB) | 26 |  | >90 |
| Social-Communication impairment (SCI) | 88 |  | 74 |

Developmental, adaptive, and behavioral assessment results for a female in the 6–10 year age range with the SYTI p.Asp366Glu (D366E) variant. Developmental functioning was assessed using the Mullen Scales of Early Learning (MSEL), adaptive functioning using the Adaptive Behavior Assessment System, Third Edition (ABAS-3), and behavioral features using standardized caregiver-reported instruments, including the Child Behavior Checklist (CBCL) and Social Responsiveness Scale–Second Edition (SRS-2). Scores are reported as standard scores, T-scores, scaled scores, and developmental age equivalents, as appropriate. Interpretive ranges are based on age-adjusted normative data from each instrument's manual.

**Table 2.**
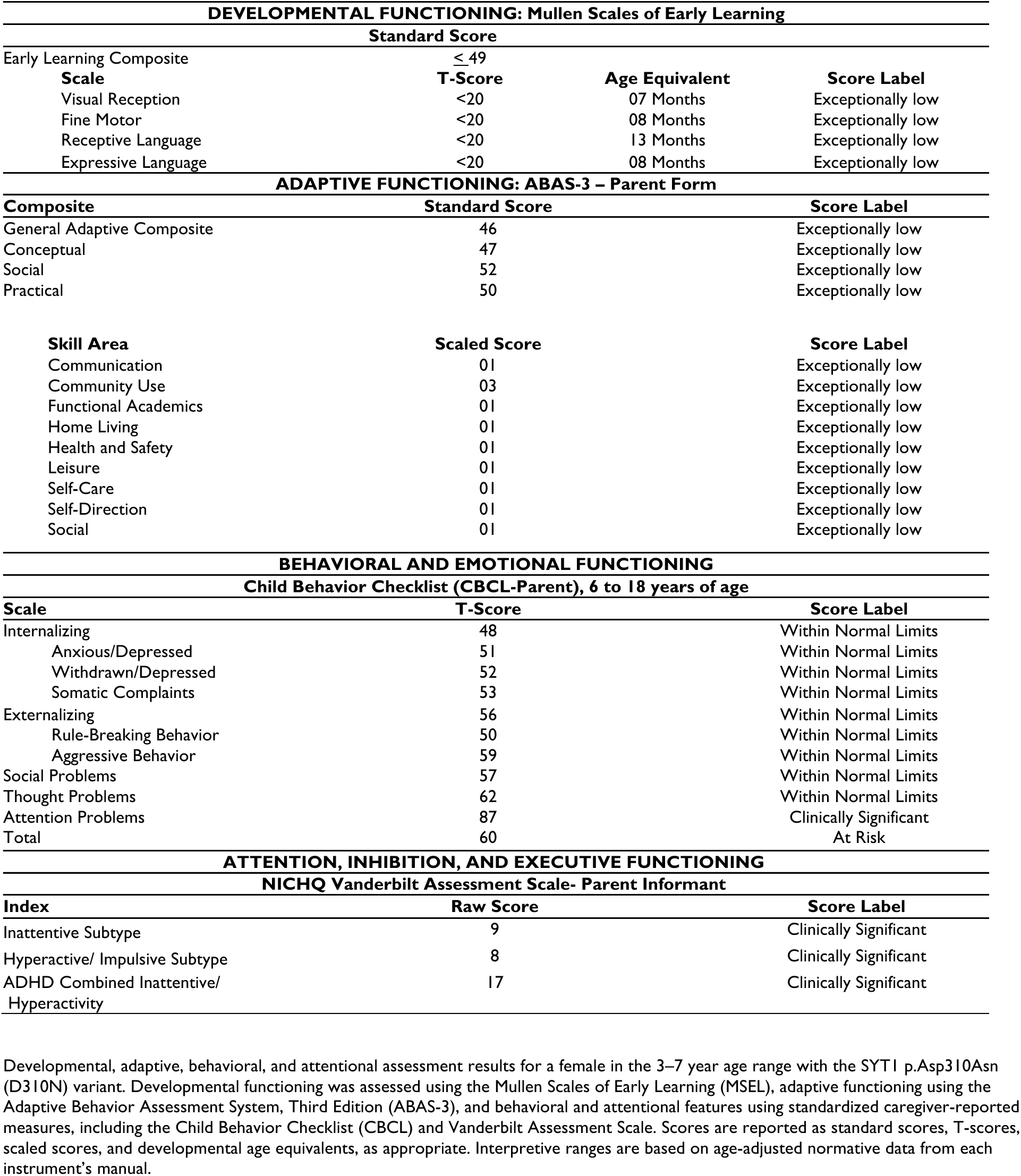
Developmental, adaptive, behavioral, and attentional assessment results for a female in the 3–7 year age range with the SYT1 p.Asp310Asn (D310N) variant.

The individual with the p.Asp366Glu (D366E) variant (female, age 6–10 years) exhibited profound developmental impairment on the Mullen Scales of Early Learning, with an Early Learning Composite score below the measurable range (<49) and developmental age equivalents clustered within early infancy across visual reception, language, and motor domains (approximately 7–16 months). Adaptive functioning, assessed using the ABAS-3, was uniformly impaired, with exceptionally low scores across conceptual, social, and practical domains (General Adaptive Composite = 50). Behavioral assessment identified clinically significant elevations in attention problems, emotional reactivity, social withdrawal, and externalizing behaviors, with marked social communication deficits and restricted and repetitive behaviors on autism-related measures.

The individual with the p.Asp310Asn (D310N) variant (female, age 3–7 years) also demonstrated severe global developmental delay, with a Mullen Early Learning Composite score below the measurable range and developmental age equivalents extending modestly higher across domains (approximately 13–23 months) relative to chronological age. Adaptive functioning on the ABAS-3 was profoundly impaired across conceptual, social, and practical domains (General Adaptive Composite = 46), with limited relative strengths observed in home living and motor skill areas. Behavioral findings in this individual were more circumscribed, with clinically significant attentional symptoms identified on the CBCL and Vanderbilt Assessment Scale and fewer elevations across other behavioral and emotional domains.

Standardized score ranges and interpretive descriptors for all developmental, adaptive, and behavioral measures are summarized in **Supplemental Table 1**.

### Differential structural perturbations in SYT1 C2B domain reveal greater Ca²⁺-binding loop destabilization by D310N than D366E

The clinical observations suggest different severity of symptoms in *Syt1^D310N^* and *Syt1^D366E^*patients. To better understand these differences, we performed molecular dynamics simulations and revealed distinct structural and dynamic consequences for the D310N and D366E variants relative to wild-type *SYT1* C2B domain. D310N exhibited the largest departure from the starting structure around the Ca²⁺-binding loops whereas D366E remained closer to wild-type. The loss of negative charge at position 310 weakened the electrostatic network that normally stabilizes loop geometry and allowed greater solvent penetration into the region. In contrast, D366E preserved the residue’s negative charge and generated only modest increases in flexibility that were confined to residues in the immediate vicinity of position 366. Neither variant produced detectable instability in the distal β-strands, indicating that the effects remained largely localized rather than destabilizing the global fold. Ca²⁺ coordination was differentially affected by the two substitutions. D310N displayed frequent and reproducible disruptions in Ca²⁺-coordinating distances. D366E trajectories retained Ca²⁺-bound states similar to wild-type, whereas D310N favored misaligned and more solvent-exposed conformations. Together, these findings indicate that D310N produces a markedly greater perturbation of C2B dynamics than D366E.

### Homozygous *syt1* mutants exhibit locomotor and synaptic transmission deficits

The function of SYT1 has been extensively characterized (*2, 5, 6, 27, 49–53*). Understanding SYT1 has taken on renewed urgency with the recent discovery that *de novo SYT1* mutations cause the rare and severe neurodevelopmental disorder Baker-Gordon syndrome (BAGOS) (*8, 9*). To date, no animal model of BAGOS has been available. In parallel with clinical evaluation of two affected individuals, we therefore sought to generate *Drosophila* models of BAGOS. Using CRISPR–Cas9, we created *Drosophila* mutants harboring variants equivalent to the patient mutations: a *dsyt1*^D362N^ allele corresponding to the human D310N variant and a *dsyt1*^D418E^ allele corresponding to the human D366E variant (**Figure 1A–B**). Throughout this work, these mutants are referred to as the *Drosophila* mutants or, in abbreviated form, *dDN* and *dDE*, respectively.

**Figure 1.**
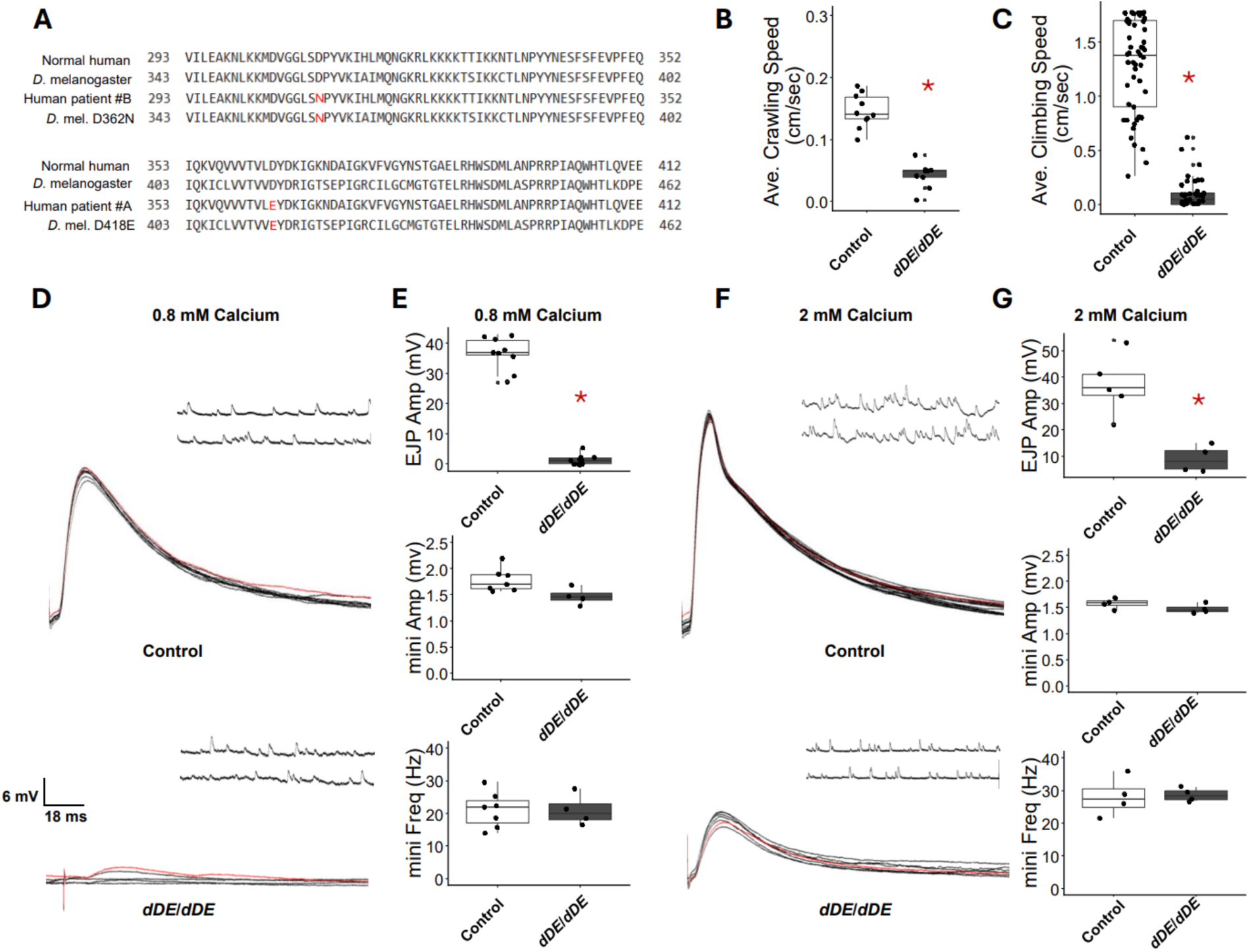
Homozygous *syt1* mutants exhibit severe defects in locomotion and evoked synaptic transmission. (A) Amino acid alignment comparing wild-type human SYT1, wild-type *Drosophila* Syt1, the human patient variants, and the corresponding *Drosophila* mutant equivalents (*dsyt1^D362N^*, *dDN* and *d*syt1^D418E^, *dDE*). (B) Larval crawling speeds of homozygous mutants (*dDE/dDE*) and controls. (C) Adult climbing speeds for homozygous mutants (*dDE/dDE*) and controls. (D) Representative EJP and miniature (mini) traces recorded in 0.8 mM Ca²⁺ HL-3 saline. (E) Quantification of mean EJP amplitude, mean mini amplitude, and mean mini frequency at 0.8 mM Ca²⁺. (F) Representative EJP and mini traces recorded in 2 mM Ca²⁺ HL-3 saline. (G) Quantification of mean EJP amplitude, mean mini amplitude, and mean mini frequency at 2 mM Ca²⁺.

Patients with BAGOS exhibit a broad spectrum of clinical features (*8–11, 54*), but motor impairment is one of the most consistently observed symptoms. To determine whether our *Drosophila* mutants display analogous locomotive defects, we performed two behavioral assays: larval crawling (**Figure 1B**) and adult climbing (**Figure 1C**). Initial testing of homozygous animals revealed that the *dDN/dDN* mutation is homozygous lethal. The *dDE/dDE* mutation was semi-lethal as homozygous at room temperature but produced viable adults when maintained at 18 °C. These *dDE/dDE* larvae exhibited markedly reduced crawling, and adult *dDE/dDE* flies showed profoundly impaired climbing ability (p = 0.0002 and p = 1.1 × 10⁻¹⁶, respectively), indicating severe locomotor dysfunction.

To determine whether the locomotor defects arise from impaired synaptic transmission, we measured evoked excitatory junction potentials (EJPs) at the larval neuromuscular junction (NMJ). In 0.8 mM Ca²⁺ HL-3, *dDE/dDE* larvae showed near-complete failure of synaptic transmission, with frequent evoked failures and occasional responses of very small amplitude, averaging approximately 2 mV (**Figure 1D–E**). Control larvae exhibited mean EJP amplitudes of ∼36 mV, corresponding to an ∼94% reduction in quantal content in the mutant. Miniature end-plate potential (mEPP) amplitude and frequency were unchanged (**Figure 1D–E**). This profound reduction in evoked release is consistent with the established role of Syt1 as a Ca²⁺-dependent exocytotic trigger.

Increasing extracellular Ca²⁺ can partially rescue synaptic transmission in *syt1* mutants (*50, 51, 55*) and in BAGOS variant-expressing neurons (*9, 12*). When we raised the HL-3 Ca²⁺ concentration to 2 mM, *dDE/dDE* larvae exhibited complete elimination of evoked failures (**Figure 1F and G**) and produced EJPs with mean amplitudes of approximately 10 mV. Control EJPs averaged ∼41 mV, a value likely approaching the reversal-potential ceiling. Although mutant responses remained significantly smaller than controls (p = 0.0024), the marked increase in EJP amplitude demonstrates a substantial Ca²⁺-dependent improvement in evoked release in homozygous mutants.

### Heterozygous *syt1* mutants exhibit climbing defects and result in seizure-like behavior

Because all reported BAGOS patients are heterozygous for *SYT1* variants, a more accurate genetic model should likewise be heterozygous animals. We therefore performed the same behavioral and electrophysiological assays on heterozygous mutants to learn whether these two variants will show behavioral or physiological defects consistent with clinical observations and molecular dynamic simulations. During the larval stage, only *dDN/+* larvae exhibited significantly reduced crawling speeds relative to controls (p = 0.04), and *dDE/+* larval crawling was similar to wildtype controls. In adults, both *dDE/+* and *dDN/+* flies displayed markedly reduced climbing speeds compared with controls (Figure 2B; p = 2.7 × 10⁻³³ and p = 0.00005, respectively). Notably, the climbing defect is more severe in the *dDN/+* compared to *dDE/+* (**Supplementary videos 1 and 2**).

**Figure 2.**
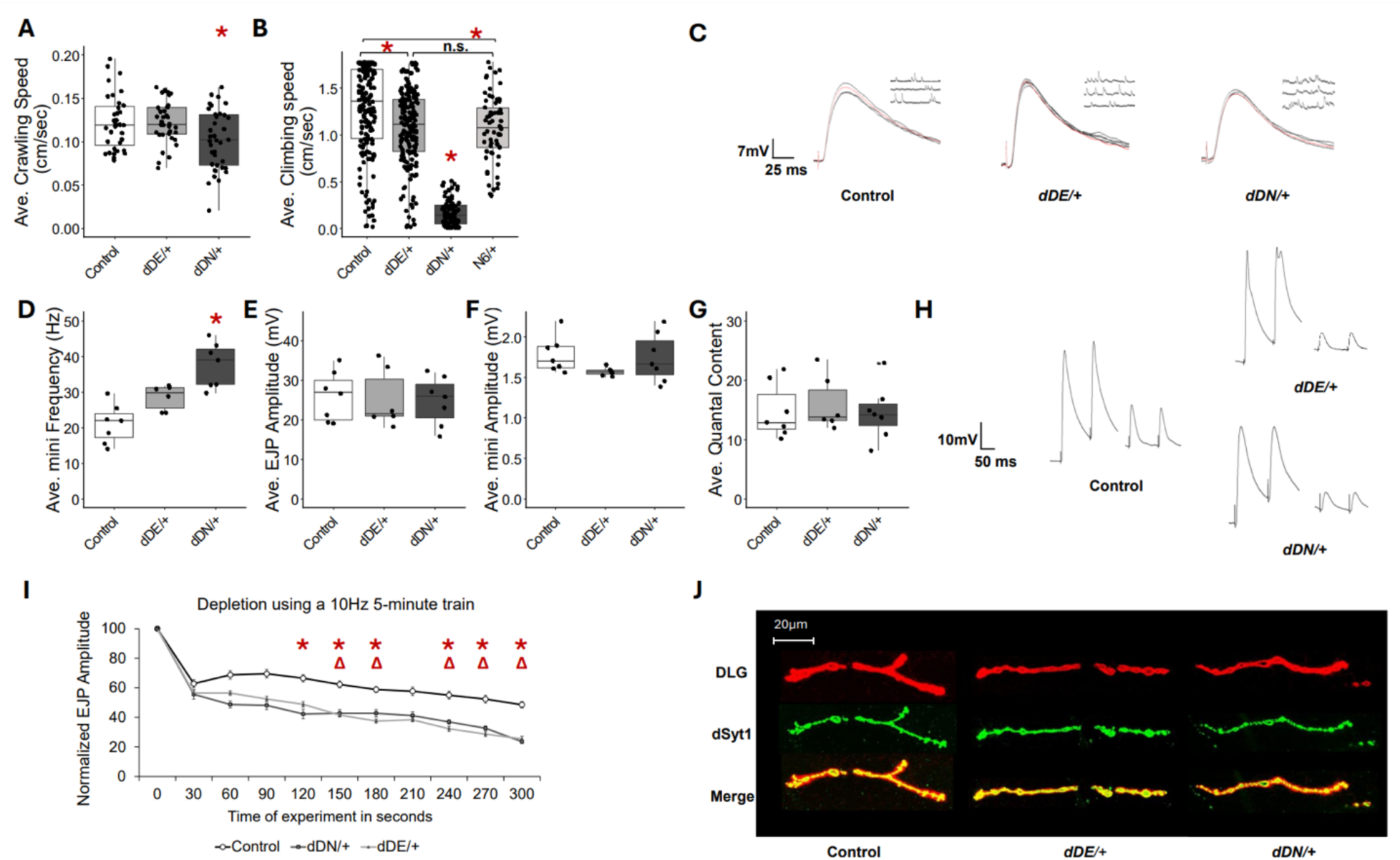
Heterozygous *syt1* mutants exhibit adult locomotor deficits and impaired SV endocytosis. **(A)** Average larval crawling speeds for each genotype (control, *dDE/+; dDN/+*). **(B)** Average adult climbing speeds at day 5; asterisks indicate *p* < 0.05. Fly genotypes are wildtype control, *dDE/+; dDN/+,* and *dsyt1 null/+ (dsyt1^N6^/+)*. **(C)** Representative EJP and miniature (mini) traces recorded from heterozygous mutants and controls. **(D)** Average miniature frequency across genotypes. **(E)** Average EJP amplitudes. **(F)** Average mEPP amplitudes. **(G)** Average quantal content, calculated as mean EJP amplitude divided by mean mEPP amplitude. **(H)** Representative traces showing the first two and last two responses of a 5-minute, 10-Hz stimulation train (1.5 mM Ca²⁺). **(I)** Normalized EJP amplitudes across the full 5-minute, 10-Hz stimulation paradigm. Delta represents statistical significance between C155>DE and C155/+ while asterisks show significance between C155>DN and C155/+ **(J)** NMJ immunostaining showing Syt1 (red) and DLG (green) in control and mutant larvae.

Both mutants displayed seizure-like behaviors during the climbing experiments upon being forced to the bottom of the graduated cylinder. To further observe this behavior, we performed the bang-sensitivity assay and video recorded their behaviors. This seizure-like behavior was observed as the flies struggling to upright themselves and jumping around spastically (**Supplemental videos 3-5**). This phenotype was more pervasive and longer lasting in the *dDN/+* heterozygous mutants. Upon further examination of the climbing movements, it was noted that the *dDN/+* mutants exhibit an ataxia-like uncoordinated walking gait which further contributed to their reduced climbing speeds (**Supplemental video 4**).

Previous studies of human patients and cell cultures support the notion that *Syt1* variants are dominant negative (*8, 12*). To test this idea, we examined *Drosophila null/+* (*dsyt1^N6^/+*) in our climbing and bang-sensitive tests and showed that *null/+* was slightly defective in climbing compared to controls but not different from *dDE/+* (Figure 2B). *dDN/+* was more severe in climbing defects among these genotypes. In bang-sensitivity tests, only two *SYT1* variants, but not *null/+* flies, showed a significantly higher percentage of seizure-like behavior (**Table 3, Supplementary video 6**). These behavioral data together argue that *dsyt1 null/+* is partially haploinsufficiency depending on the type of behaviors tested and that *SYT1* variants are dominant negative.

**Table 3.** Seizure-like behavior occurs more frequently in heterozygous mutants.

| Bang-sensitive Assay Behavioral Characteristics |  |  |  |
| --- | --- | --- | --- |
| Genotype | Seizure-like Activity | Percentage | Ave. Duration of Activity |
| Control | 13/103 | 12.6% | 1.8 sec +/- 0.74 sec |
| *dDE/+* | 42/131 * | 32% | 4.3 sec +/- 0.8 sec |
| *dDN/+* | 27/76 * | 35.5% | 4.8 sec +/- 1.5 sec |
| *Null/+* | 29/126 | 23% | 3 sec +/- 0.33 sec |

Asterisks represent statistical significance between controls using a Chi-Squared test. P-values are 0.0004 and 0.0006, respectively. Data are presented as Mean +/- SEM.

### Activity-dependent synaptic vesicle (SV) recycling is impaired in heterozygous *Drosophila syt1* mutants

Given the locomotor impairments observed in the heterozygotes, we anticipated corresponding defects in synaptic physiology. Based on prior studies, the expected phenotype would be a dominant reduction in Ca²⁺-evoked exocytosis (*8, 9, 12*). Unexpectedly, however, excitatory junction potential (EJP) amplitudes in both *dDE/+* and *dDN/+* larvae were indistinguishable from controls (**Figure 2D**). Quantal content and miniature (mini) amplitudes were also unchanged. Notably, *dDN/+* larvae exhibited a significantly elevated mini frequency (**Figure 2E–F**; p = 0.0002). These findings indicate that Ca²⁺-evoked exocytosis is preserved in heterozygous mutants, despite the clear behavioral deficits.

SYT1 contributes to both exocytosis and endocytosis (*49, 56, 57*), prompting us to examine whether synaptic vesicle (SV) endocytosis was impaired in the heterozygous mutants. To assess this, we challenged the endocytic machinery with a 5-min, 10-Hz stimulation train, a paradigm widely used to probe endocytic/recycling capacity at the *Drosophila* NMJ (*38, 58, 59*). Both *dDE/+* and *dDN/+* larvae exhibited normal EJP amplitudes at the beginning of the train but showed significantly reduced amplitudes by the end (**Figure 2H–I**; p = 0.022 and p = 0.0078, respectively). Differences between mutants and controls emerged within the first two minutes of stimulation, indicating an early failure to sustain release. These findings suggest that both heterozygous mutants exhibit deficits in SV endocytosis. Immunostaining confirmed that SYT1 is appropriately localized to the neuromuscular synapse in both variants (**Figure 2J**) and appropriately localized in the CNS (**Supplemental Figure 1**). Synaptic morphology was not affected in these mutants (**Supplementary Figure 2**).

### Heterozygous ***syt1*** mutants are defective in long-term memory

BAGOS patients consistently exhibit intellectual disability (*8–10, 54*), including the two children examined in this study (**Tables 1-3**). To assess whether the heterozygous *Drosophila* mutants show analogous cognitive impairments, we subjected *dDE/+* and *dDN/+* flies to the established courtship suppression learning and memory paradigm (*44, 60, 61*). Depending on the duration of training, courtship conditioning can be used to evaluate short-term or long-term memory (*60, 62*). In this assay, male flies are trained through repeated rejection by mated females and are subsequently tested for short-term memory one hour later and for long-term memory 24 hours after the onset of training. Learning and short-term memory performance were normal in both heterozygous mutants (**Figure 3A**). In contrast, long-term memory was profoundly impaired in both *dDE/+* and *dDN/+* flies (**Figure 3B**). One point to note is that the *dDN/+* flies had significantly reduced baseline courtship compared to controls and *dDE/+*flies (two tailed t-test p-value= 1.8×10^-8^) and may present complications when identifying a reduction in courtship.

**Figure 3.**
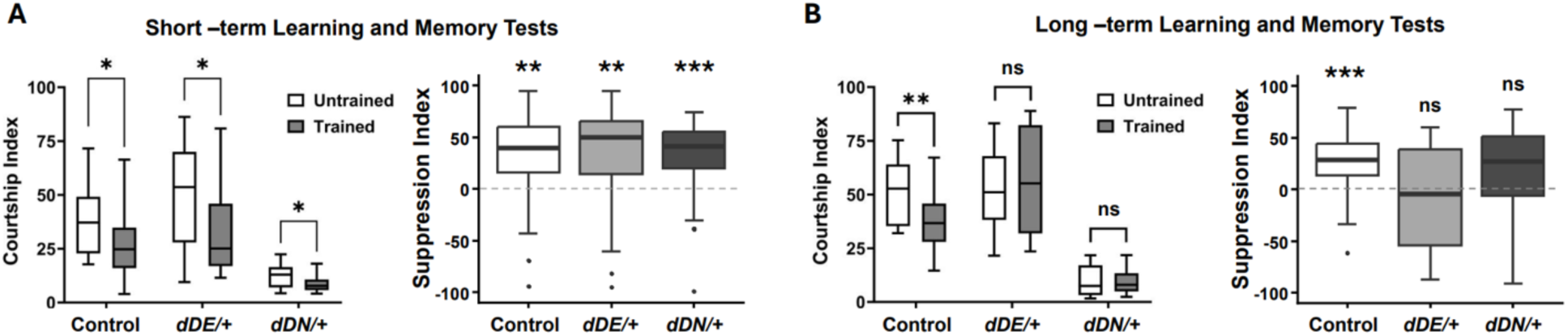
Heterozygous mutants are defective in long-term memory. **(A)** Left, Courtship index values for male flies in untrained and trained groups for each genotype. Flies were trained for 1h and tested for short-term memory 1h after the end of training. Sample size (untrained:trained) is (38:31) for CS/+, (36:34) for *dDE/*+, and (36:33) for *dDN*/*+*. Right, Suppression index. *p<0.05, **p<0.01. **(B)** Left, Courtship index values for male flies in untrained and trained groups for each genotype. Flies were trained for 6h and tested for long-term memory 24h after the onset of training. Sample size (untrained:trained) is (32:29) for CS/+, (30:31) for *dDE/+*, and (31:29) for *dDN/+*. Right, Suppression index. **p<0.01, ***p<0.001, n.s.= not significant.

### Neuronal expression of human *SYT1* variants recapitulates locomotor and synaptic defects of heterozygous *syt1* mutants

Our heterozygous *Drosophila syt1* mutants provide a valuable genetic model of BAGOS, but this approach limits the ability to dissect cell type–specific and circuit-level effects of *SYT1* variants. To enable such studies, we generated humanized transgenic flies expressing UAS-versions of the two human variants using human *SYT1* DNA under the control of Gal4 (*63*). When UAS-D310N or UAS-D366E was expressed pan-neuronally under C155 (Elav)-Gal4, both genotypes showed significantly reduced larval crawling speeds compared with controls (p = 0.0002 and p = 0.008, respectively), and adults exhibited markedly impaired climbing performance (**Figure 4A–B**; p = 1.3 × 10⁻³¹ and 2.6 × 10⁻²⁸, respectively). Consistent with our CRISPR-generated *Drosophila* heterozygous *syt1* mutants, the UAS-D310N variant produced more severe phenotypes than UAS-D366E.

**Figure 4.**
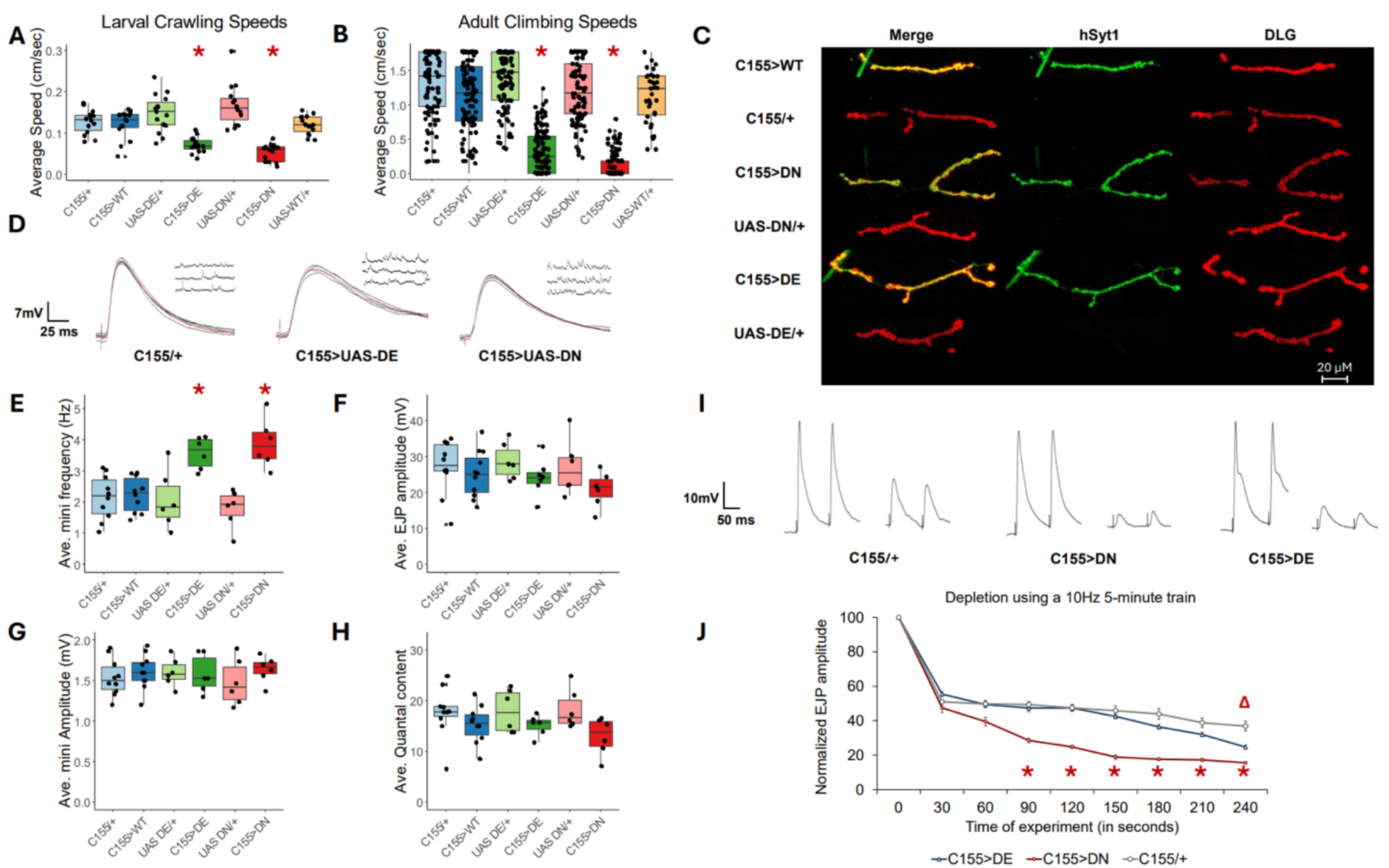
Pan neuronal expression of human *SYT1* variants slows locomotion and impairs SV endocytosis. In this set of experiments, the pan neuronal Gal4 (C155) was used to drive the expression of UAS-DE or UAS-DN variants as well as the wildtype human UAS-SYT1 in neurons. Larval crawling, adult fly climbing, larval NMJ synaptic physiology, and larval NMJ morphology were investigated. (A) Larval crawling speeds for each genotype. (B) Adult climbing speeds. (C) NMJ immunostaining of larval muscle 4 in abdominal segments A2 or A3, showing synaptic localization of wild-type and mutant human SYT1. (D) Representative EJP and miniature (mini) traces from UAS mutants and controls. (E) Average miniature event frequency. (F) Average EJP amplitudes. (G) Average mini amplitudes. (H) Average quantal content. (I) Representative traces showing the first two and last two responses during a 5-minute stimulation train. (J) Normalized EJP amplitudes across the entire 5-minute stimulus paradigm. Delta represents statistical significance between C155>DE and C155/+ while asterisks show significance between C155>DN and C155/+

Using an antibody specific to human SYT1, we confirmed that wild-type and mutant human SYT1 proteins localize appropriately to the synapse; fluorescence was observed in all genotypes expressing UAS constructs, including C155>UAS-WT, C155>UAS-D310N, and C155>UAS-D366E, but absent in control larvae, C155/+ and UAS-syt1/+ (**Figure 4C**). Staining for endogenous *Drosophila* Syt1 further confirms the proper synaptic localization at the NMJ (**Supplemental Figure 3**) and in the CNS (**Supplemental Figures 4 and 5**) and shows that synaptic morphology appeared normal (**Supplementary Figure 3**). Thus, human mutant SYT1 as well as the native *Drosophila* Syt1 protein traffic normally.

**Figure 5.**
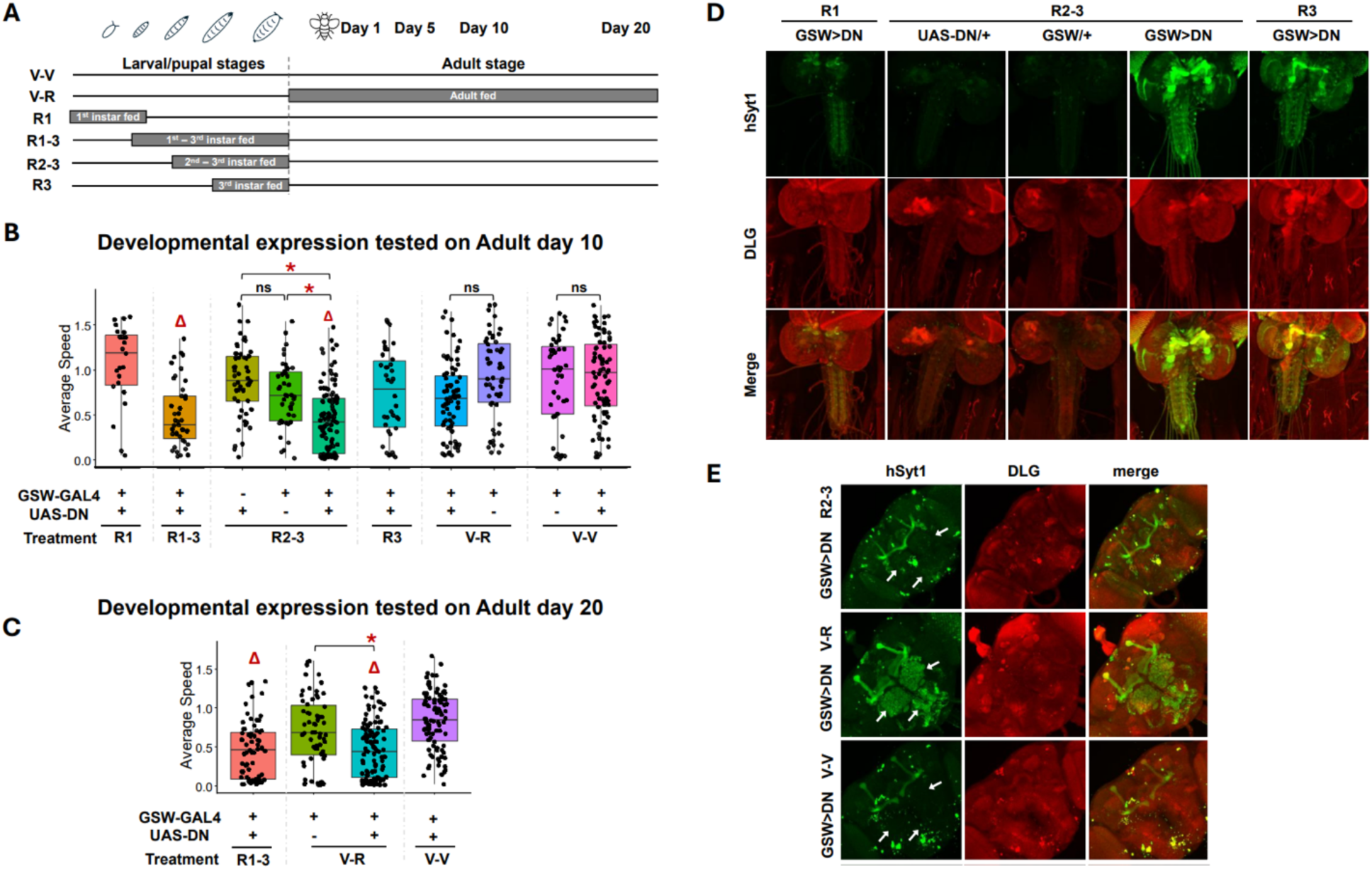
Early larval and pupal expression of mutant *SYT1* is sufficient to induce locomotive defects. In this set of experiments, the DN variant was induced to express in neurons during defined developmental stages using a drug-inducible Elav-Gal4 (called GeneSwitch). The experimental flies along with control groups were then tested for climbing and their brains examined for human SYT1 expression. **(A)** Schematic of the GeneSwitch RU486-feeding paradigm used to induce stage-specific expression of UAS-D310N. V-V: food contained vehicle; V-R: fed with RU-containing food during adult stage; R1: fed with RU-containing food during first instar stage; R1-3: fed with RU-containing food during first instar to third instar stages; R2-3: fed with RU-containing food during second instar to third instar stages; R3: fed with RU-containing food during third instar stage. **(B)** Adult climbing performed on day 10 across all experimental and control groups demonstrates persistent locomotor impairment following early developmental expression of the mutant variant. Note the difference between R2-3 and R3 groups. Asterisks represent significance between genotypes and deltas represent statistical significance between vehicle treated GSW>DN controls. **(C)** Climbing behavior tested on day 20 shows that continuous and prolonged adult expression of DN causes climbing defects to a level similar to those expressed in R1-3 group. Asterisks represent significance between genotypes, and deltas represent significance between vehicle treated GSW>DN controls. **(D)** Immunocytochemistry shows the presence or absence of human SYT1 (green) expression in different experimental and control larvae. The larval CNS preparation was also stained for DLG (red) and performed on 3^rd^ instar stages. **(E)** Adult fly brains stained for hSYT1 (green) and DLG (red) in the R2-3 (left panel), V-R condition (middle panel), and of the V-V condition (right panel) from the GSW>DN genotypes. Note that R2-3 and V-V brains display no detectable SYT1 in the neuropil areas in adult brains (left panel, see arrows) compared to SYT1 in the neuropil areas in V-R brains (see arrows). This antibody non-specifically stains the mushroom body and shows ‘junk’ signals elsewhere.

Next, we examined the synaptic physiology of the pan neuronal Gal4-driven human variants and found that they closely phenocopy the defects observed in the heterozygous *Drosophila* mutants. Specifically, both C155>UAS-D310N and C155>UAS-D366E larvae exhibited significantly elevated miniature event frequencies relative to controls (p = 0.005 and p = 0.003, respectively), no change in EJP amplitude, mini amplitude and quantal content (**Figure 4D–E**). In addition, both variants showed clear defects in SV recycling during sustained stimulation while maintaining normal basal transmitter release (p = 0.02 and p = 0.05, respectively) (**Figure 4D–J**).

### Temporal induction of the *SYT1 D310N* variant identifies a critical developmental window for the onset of climbing defects

While SYT1 has been extensively studied for its roles in synaptic transmission, few studies have addressed its developmental role. The availability of UAS-*SYT1* variants enabled us to investigate whether specific developmental windows are critical for the emergence of disease-related phenotypes. To do this, we employed the GeneSwitch inducible Gal4 system (*34, 35*) to drive expression of UAS-D310N at defined developmental stages using RU486-dependent activation. In our feeding paradigm (**Figure 5A**), V-V flies were maintained on vehicle-treated food for the entire experimental period and served as controls. V-R flies received RU486 only during adulthood. The R1 group received RU486 from embryogenesis through late first instar; the R1–3 group was fed RU486 from first instar through pupation; the R2–3 group received RU486 beginning in second instar and continuing through pupation; and the R3 group was exposed to RU486 from third instar until pupation. After the designated exposure periods, all flies were transferred to vehicle-treated food.

Our data show that brief induction of UAS-D310N during either the embryonic-to–late first instar stage (R1) or during the third instar–to–pupal transition (R3) does not affect adult climbing ability (**Figure 5B**). In contrast, induction of D310N expression from second instar through pupation (R2–3) is sufficient to significantly impair adult climbing, suggesting that the critical period resides in 2^nd^ instar stage. We stained 3^rd^ instar larvae under the R1, R2-3 and R3 conditions to verify that mutant SYT1 was expressed under these acute RU exposed conditions, and indeed we observed strong expression of the mutant SYT1 in the R2-3 and R3 conditions and less expression under R1 conditions (**Figure 5C**). However, SYT1 in R2-3 is undetectable in adult brains on day 5 (**Figure 5D**). Interestingly, D310N expression restricted to adulthood (V-R) does not alter climbing performance on days 5 or 10, despite strongly mutant SYT1 protein expressed in adult brains (**Figure 5D**). These behavioral and immunocytochemical observations show that acute expression of mutant SYT1 protein during early development produces persistent locomotor defects even though the mutant protein is no longer detectable in adult flies. Equally important, adult expression of mutant SYT1 is insufficient to induce climbing defects when tested on or before day 10.

We then wanted to know if prolonged expression of D310N would be able to affect climbing. When tested on 20 days, V-R flies indeed showed climbing defects (**Figure 5E**). These results suggest that just D310N expression alone in adult flies is insufficient to produce climbing defects prior to 20 days, but longer expression duration is needed to override or corrupt the existing climbing circuits. V-R flies were as poor climbers as the flies from the R1-3 group. As pointed out by Bains and colleagues (*64*), and based on the findings from the Broadie group (*65*), there appears to be multiple ‘critical periods’ for the onset of behavioral plasticity. Our data suggests that climbing defects can be more readily induced during mid larval stage and reluctantly in adults. Together, these results support the notion that BAGOS is fundamentally a developmental disorder and further suggest that acute synaptic dysfunction alone is unlikely to account fully for the disease phenotype.

### Neuron-subset–specific expression identifies diverse contributors to seizure-like activities and climbing defects

The Gal4-UAS method is versatile in that it allows us to dissect the contribution of specific cell types in this disease model. To this end, we expressed UAS-*SYT1* variants using Gal4 drivers that cover subset types of neurons, including cholinergic neurons (Chat-Gal4), GABAergic neurons (Gad1-Gal4), monoamine neurons (Ddc-Gal4), motoneurons (Ok6-Gal4), and glutamatergic neurons (vGlut-Gal4). For UAS-D310N, we found that Chat-Gal4 and GAD1-Gal4 drivers were able to induce severely slow climbing speeds compared to both parental controls (p= 4.7×10-15 and 2.9×10-9, respectively) (**Figure 6A**), and that OK6-Gal4 expression also significantly reduces climbing speeds in DN but not DE (and to a lesser degree than Chat-Gal4 and GAD1-Gal4) (p=0.008). Surprisingly, vGlut-Gal4-driven D310N expression did not have a detectable effect on climbing. For UAS-D366E, we found that only the Chat-Gal4 expression resulted in significantly slower climbing compared to both parental controls (p = 0.009), while Ddc-Gal4 expression resulted in significantly slower climbing compared to Ddc-GAL4/+ (p=0.004) control but not to the UAS-DE/+ control (**Figure 6B**).

**Figure 6.**
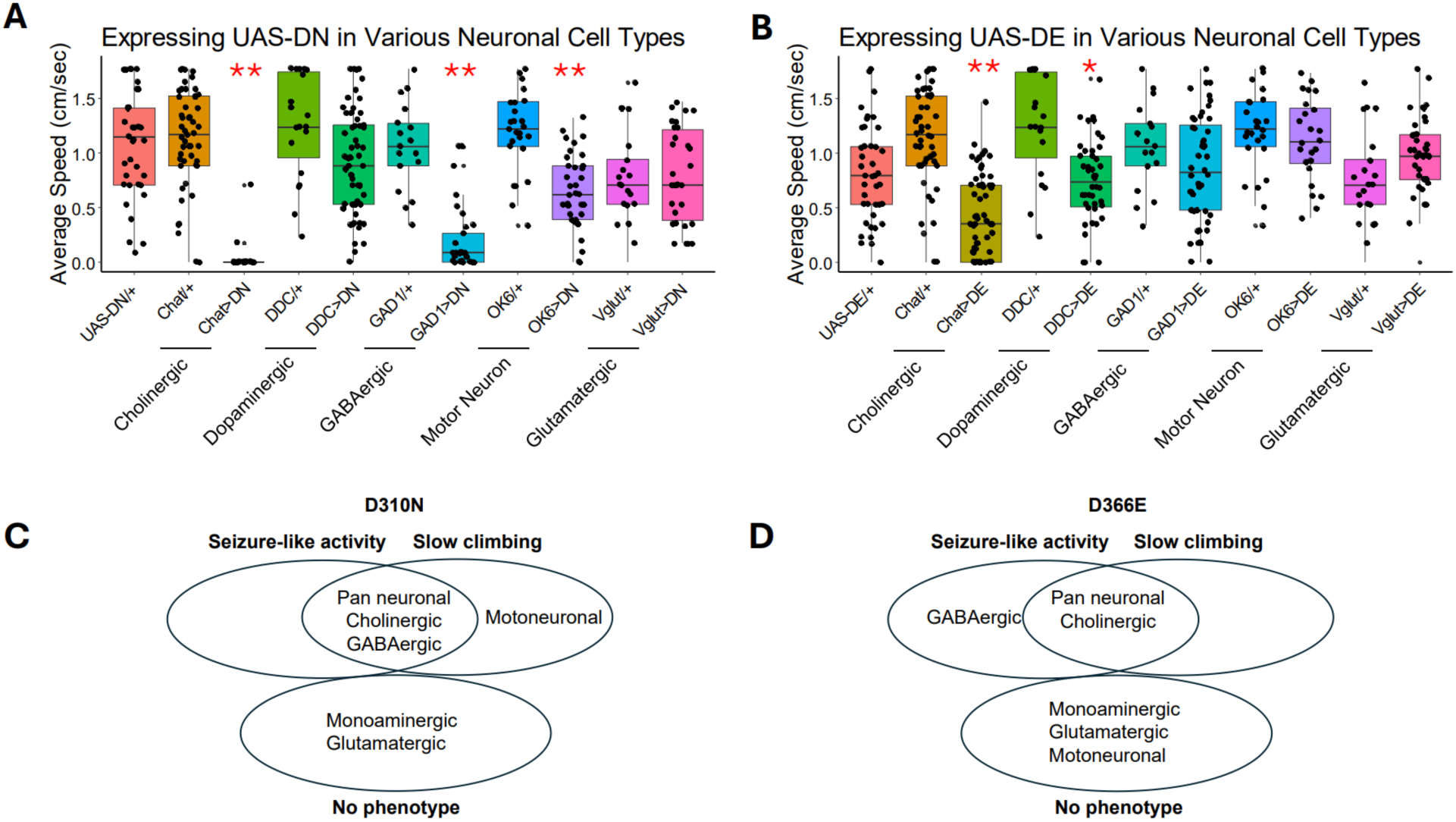
Cholinergic neurons are primary drivers of the impaired climbing phenotype whereas expression of mutant *SYT1* in cholinergic and GABAergic neurons induces seizure-like behavior. (A) Average climbing speeds of 5-day-old adult flies expressing UAS-D310N under various tissue-specific Gal4 drivers. From left to right: control, cholinergic (Chat-Gal4), dopaminergic/monoaminergic (Ddc-Gal4), GABAergic (Gad1-Gal4), motor neuron (Ok6-Gal4), and glutamatergic (vGlut-Gal4). UAS-D310N/+ and Gal4/+ serve as controls. ** indicates significant differences when compared to both controls. (B) Average climbing speeds of flies expressing UAS-D366E under the same set of Gal4 drivers. UAS-D366E/+ and Gal4/+ serve as control. ** indicates significant differences when compared to both controls. * indicates a significant difference when compared to Gal4/+, but not to UAS-D366E/+. (C) Venn diagram summary indicating whether each Gal4 driver produced seizure-like activity and/or slowed climbing when driving expression of the D310N variant. (D) Venn diagram summary indicating whether each Gal4 driver produced seizure-like activity and/or slowed climbing when driving expression of the D366E variant.

The bang-sensitivity assay uses forceful taps to illicit uncoordinated movements in flies. The first part of the climbing assay is comparable to the bang-sensitivity assay. We found that expression of D310N or D366E in either Chat or Gad1-Gal4 was sufficient to induce seizure-like activities immediately after banging down during the climbing tests (**Figure 6C and D**). The Gad1-Gal4 data reveal that climbing defects and seizure-like activity depend on the *SYT1* variant being expressed. Surprisingly, expression of *SYT1* variants in motoneurons only affected D310N flies whereas both variants in glutamatergic neurons did not have detectable effects on fly climbing or seizure-like activity. These findings suggest that BAGOS is not a result of simple synaptic dysfunction, but rather of disordered neural networks.

## Discussion

In this study, we employed a reverse-translation strategy, using human clinical observations to generate the first animal models of Baker-Gordon Syndrome (BAGOS) carrying heterozygous mutations identical to patient genetic variants. Using both CRISPR-engineered *Drosophila* mutants and humanized UAS transgenes, we examined how two patient-derived *SYT1* variants, D310N and D366E, alter synaptic physiology, locomotion, memory, circuit function, and developmental timing. These *in vivo* findings reveal mechanisms that cannot be inferred from cultured neurons alone and offer new insight into how *SYT1* dysfunction disrupts brain development and behavior.

### Synaptic physiology

Syt1 is well established as the principal Ca²⁺ sensor for synchronous neurotransmitter release and a contributor to synaptic vesicle endocytosis (*7*). The C2B domain contains Ca²⁺-binding loops essential for Ca²⁺-dependent lipid penetration and rapid fusion (*7, 52, 53, 66, 67*). It interacts with SNARE complexes in ways that regulate fusion clamping and fusion triggering (*52, 68*).

The C2A domain also binds Ca²⁺ and influences release kinetics and cooperativity (*69*). Since the identification of *SYT1* as the gene underlying BAGOS, cultured-neuron studies have shown that most variants traffic normally to synapses but display dominant negative effects on exocytosis, endocytosis, or both (*8, 9, 12, 54*). Some variants, such as I368T, even produce mixed phenotypes, with reduced Ca²⁺-evoked release but enhanced SV recycling (*8*).

Our findings show that the *Drosophila* equivalents of D310N and D366E produce severe physiological consequences *in vivo. dsyt1*^D310N^ homozygotes die during embryogenesis, consistent with a strong loss-of-function or dominant-negative mechanism.

*dsyt1*^D366E^ homozygotes survive to larval stages and, when raised at 18 °C, reach adulthood, similar to *syt1* nulls (*27*). Electrophysiological analysis of *syt1* D366E homozygous larvae demonstrated markedly diminished evoked transmission, high failure rates, and more than 90 percent reduction in EJP amplitude. Raising extracellular Ca²⁺ partially improved the response, consistent with work showing that increased Ca²⁺ can compensate for impaired Syt1-mediated triggering (*9, 12, 50, 51, 55*). Although D310N homozygotes could not be assessed because of early lethality, the clinical severity and behavioral phenotypes associated with D310N are consistent with a more severe impairment of synaptic function.

An unexpected result emerged when examining heterozygotes. Despite the clear clinical and behavioral defects in BAGOS patients, and despite dominant negative effects documented in cultured neurons (*8, 9, 12*), we detected no reduction in single-pulse Ca²⁺-evoked release in heterozygous D310N or heterozygous D366E animals. However, both variants produced robust deficits during sustained and repetitive activity, revealing accelerated synaptic depression due to impaired SV recycling. This pattern was confirmed when expressing humanized versions of the variants pan-neuronally. It is possible that developmental compensation or circuit-level regulation in intact animals preserves basal release, whereas high-frequency or sustained activity unmasks the recycling deficits. These findings highlight the importance of studying synaptic function *in vivo*, where the balance of protein expression, network-level context, and developmental constraints differ markedly from cultured systems.

### Climbing defects, seizure-like activity, and memory deficit

Motor impairment is among the most consistent features reported in BAGOS patients (*8, 10, 54*). Negative geotaxis in *Drosophila* provides a robust and widely used behavioral assay for aging studies (*28, 29*) and for detecting neurological dysfunction across models of Parkinson’s disease (*70*), Alzheimer’s disease (*71*), ALS (*72*), and autism-associated gene variants (*30, 73*). Both heterozygous D310N and D366E flies exhibited substantial climbing impairments that align with human motor phenotypes. Pan-neuronal expression of the UAS transgenes further recapitulated these defects, providing a versatile framework for probing cell-specific mechanisms.

Seizure-like activity was also more severe in D310N-expressing flies, a finding consistent with the variant’s stronger SV impairment. Similar locomotor instability has been described in mutants that affect release probability, synaptic vesicle replenishment, or excitatory and inhibitory balance (*74*). These parallels suggest that BAGOS variants disrupt the coordinated activity of interneuron networks that stabilize locomotor patterns.

Long-term memory defects in both heterozygotes further echo the intellectual disabilities observed in BAGOS patients. *Drosophila* long-term memory depends on CREB-driven transcription, neuromodulation, and circuit-level plasticity (*75, 76*). Several synaptic release mutants, including *synapsin*, *rab3*, *unc-13*, and *syt1* hypomorphs, show selective impairment of long-term but not short-term memory (*77*). The preservation of short-term memory in our models suggests that basal synaptic function remains adequate for immediate behavioral performance but insufficient for stabilizing plastic changes required for long-term consolidation.

### Synaptic function, circuits, and development

A central question in BAGOS pathogenesis is whether synaptic dysfunction alone is sufficient to cause the disorder or whether circuit-level and developmental mechanisms play a central role.

Our findings strongly support the latter interpretation, indicating that BAGOS is fundamentally a developmental disorder of neural circuit assembly.

One major clue lies in the cell-type specificity of behavioral phenotypes. Expression of D366E in motoneurons did not impair climbing despite clear physiological deficits at the neuromuscular junction. This suggests that intact animals compensate for peripheral synaptic impairment and points to central circuits as the primary locus of dysfunction. In contrast, expression of either variant in cholinergic neurons-which represent key interneuron populations in the pattern-generating locomotor circuits (*78, 79*) - produced strong climbing deficits. The difference between D310N and D366E also aligns with their synaptic phenotypes. D310N, which produces more severe recycling defects, perturbs both cholinergic and GABAergic circuits. D366E affects climbing primarily through cholinergic neurons. Seizure-like activity is induced by expressing D310N or D366E in cholinergic neurons and by D310N expression in GABAergic neurons.

These findings parallel observations in disorders such as Rett syndrome and Dravet syndrome, where interneuron dysfunction is a major driver of the clinical phenotype (*80, 81*).

The GeneSwitch experiments provide strong evidence that BAGOS reflects disrupted early neural development. Transient expression of D310N during the second instar to pupal transition was sufficient to produce persistent adult locomotor deficits, and yet no D310N protein can be detected in the adult fly brain. Whereas adult-only expression, despite robust expression levels, had no measurable effect on climbing until 20 days of expression. Although the specific cellular or circuit-level changes that persist following this brief developmental expression were not directly assessed, the irreversibility of the phenotype is consistent with disruption of activity-dependent circuit maturation during a critical developmental window. This aligns with the concept of critical periods in circuit maturation, when activity-dependent refinement shapes stable connectivity patterns (*64, 82–86*). Given the demonstrated defects in SV recycling, cell-type–specific circuit vulnerability, and developmental timing sensitivity observed in this study, we hypothesize that transient *SYT1* dysfunction during this period leads to maladaptive circuit assembly or a disruption of normal circuit maturation resulting in persistent functional defects.

Together, these results suggest that BAGOS is not merely a disorder of impaired synaptic release but reflects a deeper failure of circuit formation, integration, and plasticity. Synaptic defects, particularly in recycling, likely perturb signaling within central interneuron networks during a time when precise activity patterns are essential for circuit refinement. The behavioral deficits that emerge in adulthood represent the long-lasting consequences of disrupted developmental processes rather than ongoing exocytic failures alone. Our results caution that gene therapies have to be delivered during early developmental stages to be effective.

### Linking Human BAGOS Phenotypes to Developmental Synaptic and Circuit Mechanisms

Baker–Gordon syndrome (BAGOS) illustrates how a single amino acid substitution in a presynaptic protein can profoundly influence human brain development. Affected individuals typically present in infancy or early childhood with motor delay, hypotonia, ataxia, and cognitive impairment, often accompanied by behavioral dysregulation, sleep disturbance, visual abnormalities, and impaired communication.

In our studies, both individuals with *SYT1* missense variants exhibited severe global neurodevelopmental impairment characterized by MSEL performance below the measurable range and profound adaptive dysfunction on the ABAS-3. While both cases demonstrated substantial developmental delay, differences were observed in the relative emphasis of behavioral and functional features, with the older individual showing more prominent behavioral dysregulation and autism-related symptoms and the younger individual demonstrating comparatively fewer behavioral elevations despite similarly severe adaptive impairment.

These findings are consistent with prior reports describing broad phenotypic severity and clinical heterogeneity in SYT1-associated neurodevelopmental disorder. Given the small sample size, cross-sectional design, and differences in age and developmental stage at assessment, the observed differences should be interpreted descriptively rather than as evidence of variant-specific effects. Longitudinal evaluation across larger cohorts will be necessary to clarify developmental trajectories and the relative contribution of age-dependent, environmental, and genetic factors to phenotypic variability.

Despite clinical heterogeneity, many share a recognizable constellation of features, including impaired coordination, a characteristic wide-eyed facial expression, and profound difficulties with learning and adaptive behavior. These phenotypes persist throughout the lifetime of patients (52); thus, BAGOS likely reflects disrupted developmental wiring rather than episodic synaptic failure as some medications taken by patients (e.g., 4-AP) are aimed at the synaptic failure with little success for disease phenotype amelioration.

*Drosophila* models provide a powerful complementary framework for dissecting disease mechanisms. Flies preserve core features of synaptic transmission while enabling precise control over genetic, cell-type–specific, and developmental variables. Expression of the same *SYT1* variants that impair gait and learning in children disrupts locomotor patterning, interneuron balance, and circuit maturation in the larval ventral nerve cord. Notably, the developmental period during which BAGOS symptoms emerge in humans aligns closely with a critical second-to-third instar window in flies, suggesting conserved principles of synaptic development and circuit refinement across species.

In heterozygous flies, basal Ca²⁺-evoked neurotransmitter release is preserved, yet sustained activity reveals pronounced synaptic depression due to impaired vesicle recycling. This demand-dependent failure parallels the clinical phenotype, in which deficits in coordination, learning, and behavioral regulation are most evident during dynamic or repetitive activity rather than at rest.

These findings support a model in which *SYT1* variants impose a latent vulnerability that becomes unmasked as developing circuits encounter increasing functional demand.

Variant-specific analyses further strengthen the translational relevance of the model. Our molecular dynamic simulation suggests greater perturbation of C2B by D310N than D366E. The D310N variant, associated with more severe motor and cognitive impairment in patients, produces greater locomotor deficits, broader circuit vulnerability, and more pronounced synaptic dysfunction in flies than D366E, mirroring clinical severity. Developmental timing experiments reinforce this framework: transient expression of mutant SYT1 during a restricted larval–pupal window is sufficient to induce permanent adult locomotor impairment, whereas adult-only expression has no effect. These results indicate that BAGOS arises from disrupted circuit maturation rather than ongoing synaptic failure in adulthood. Notably, the D366E variant produces only mild impairments in synaptic vesicle exocytosis in mammalian systems (*9, 87*). Baker and colleagues further demonstrated that these modest synaptic deficits correlate with comparatively mild clinical phenotypes in individuals carrying D366E, consistent with our clinical observations.

Cell-type–specific studies refine the locus of vulnerability further. Expression of mutant SYT1 in cholinergic and GABAergic interneurons is sufficient to reproduce seizure-like activity and locomotor dysfunction, whereas expression in glutamatergic neurons is not. This implicates central interneuron networks as primary drivers of disease, consistent with preserved muscle strength but impaired motor control in affected individuals and with shared principles of circuit fragility observed in other neurodevelopmental disorders.

Together, these findings support a unified model in which SYT1 variants preserve basal synaptic transmission but impair vesicle recycling under sustained activity, thereby disrupting activity-dependent processes required for normal circuit maturation during critical developmental periods. BAGOS is thus best understood as a disorder of circuit development, in which early synaptic perturbations are translated into lifelong neurological dysfunction.

## Data availability

All *Drosophila* lines, plasmids, and reagents generated in this study are available upon request, or the fly lines can be obtained from Bloomington *Drosophila* Stock Center.

Electrophysiological datasets, behavioral recordings, and analysis scripts are available from corresponding author, upon reasonable request. Clinical data from the BAGOS Natural History Study are available in de-identified form to qualified investigators following IRB and data-use approval.

## Data Availability

All data produced in the present study are available upon reasonable request to the authors

## Acknowledgments

This research was initially supported by Kamdyn Genetics, LLC established by John Ruth and family for the project *Synaptotagmin-1 Neurodevelopmental Disorder* (PI: Carney; award period 07/01/2022–12/30/2023) and by University of Missouri internal research funds (Carney).

Additional support was provided by subcontracts awarded to B.Z., D.D., and P.R.C from Genetic Autism Alliance (GAA) through the Missouri Department of Health (P.I: Paul Carney, 09/01/2023-06/01/2025; P.I.s David Arnold and Chris Lorson 06/02/2025-present). C.R. was supported in part by a University of Missouri SEC Emerging Scholars Postdoctoral Award.

We extend our deepest gratitude to the patients and families whose participation made this work possible. We thank John Ruth and family for the Kamdyn Genetics support and members of the GAA and the Center for Translational Neurogenetics, David Arnold, Andrea Delgado, De-Pei Li, Christian Lorson, Bettina Mittendorfer, John Ruth, Smita Sexena, and Kamlendra Singh, for their feedback. We are also grateful to Sophia Marchetti for assistance in organizing activities related to the Natural History Study and for organizing GAA meetings. We thank Noreen Reist for providing the *Drosophila* Syt1 antibody.

## Ethics Statement

All human studies were approved by the University of Missouri Institutional Review Board (IRB) and conducted in accordance with the Declaration of Helsinki and institutional guidelines for research involving human participants.

The BAGOS Natural History Study is registered at ClinicalTrials.gov (Identifier: NCT06399952). All procedures, including recruitment, assessment, data storage, and data handling, were approved under IRB-authorized protocols.

## Consent Statement

Written informed consent was obtained from all parents or legal guardians of participating children. Assent was obtained from participants when developmentally appropriate. Consent included permission for clinical data, de-identified videos, neurobehavioral assessments, and genetic findings to be used for research and publication.

## Author Contributions

B.Z, C.E.R., and P.R.C. conceived the project. D.J.D. generated the constructs of UAS-SYT1 variants used to produce transgenic flies and designed CRISPR-Cas9 strategies; C.E.R., J.P., B.L.H., L.M, S.D., O.A., and B. Z. performed *Drosophila* experiments. C.L.A. conducted clinical phenotyping. L.T.C coordinated the natural history studies. B.Z. supervised *Drosophila* studies except learning and memory studies, which were supervised by S.D. P.R.C performed the molecular dynamic simulations and supervised the natural history study. B.T.B conducted neurological studies of the two patients. C.E.R., J.P., P.R.C., D.J.D., S.D., and B.Z. contributed to data interpretation. B.Z., C.E.R., and P.R.C. wrote the manuscript; D.J.D. and S.D. provided editing.

## Competing Interests

The authors declare no competing financial or non-financial interests.

## Notes

### Competing Interest Statement

The authors have declared no competing interest.

### Clinical Trial

NCT06399952

### Author Declarations

Ethics Statement All human studies were approved by the University of Missouri Institutional Review Board (IRB) and conducted in accordance with the Declaration of Helsinki and institutional guidelines for research involving human participants. The BAGOS Natural History Study is registered at ClinicalTrials.gov (Identifier: NCT06399952). All procedures, including recruitment, assessment, data storage, and data handling, were approved under IRB-authorized protocols. Consent Statement Written informed consent was obtained from all parents or legal guardians of participating children. Assent was obtained from participants when developmentally appropriate. Consent included permission for clinical data, de-identified videos, neurobehavioral assessments, and genetic findings to be used for research and publication.

