## Supplementary materials for "From Bedside to Bench: *Drosophila* Models of Baker-Gordon Syndrome (BAGOS)"

Carlos E. Rivera *et al.*

**This PDF file includes:**

Supplementary Text
Figs. S1 to S5
Table S1
Movies S1 to S6

**Other Supplementary Materials for this manuscript include the following:**

Movies S1 to S6

### Supplementary Text

Molecular dynamic simulation methods in detail

The *SYT1* C2B domain structure was modeled in silico using a high-resolution crystal template, and point mutations D310N and D366E were introduced with PyMOL's mutagenesis tool while preserving backbone geometry and minimizing steric clashes. Each model was placed in a rectangular box of TIP3P water with at least 10–12 Å padding and neutralized with counterions, followed by the addition of 150 mM NaCl. Systems were parameterized with the AMBER ff14SB force field, and calcium ions were included in the canonical C2B binding pockets using established parameters. Energy minimization was performed in two stages using steepest descent and conjugate gradient algorithms. The simulated system was then gradually heated from 0 to 310 K under an NVT ensemble over 100–200 ps using a Langevin thermostat, a standard computational equilibration procedure to bring the virtual system to physiological temperature without introducing numerical instabilities. This was followed by density equilibration under NPT conditions at 1 atm for 500 ps to 1 ns with decreasing harmonic restraints on backbone atoms, employing particle mesh Ewald electrostatics and SHAKE constraints to allow a 2 fs timestep.

Production simulations were carried out under NPT conditions at 310 K and 1 atm. Three independent trajectories of 100–200 ns were generated for each system (wild-type, D310N, D366E), with coordinates saved every 10 ps. Trajectory analyses were performed using CPPTRAJ and GROMACS utilities. Backbone RMSD values were calculated relative to the minimized starting structure after alignment to the C2B β-sandwich core. Local flexibility was assessed with per-residue RMSF, and secondary structure was evaluated using DSSP. Calcium-binding loop dynamics were examined by measuring distances between coordinating residues and $Ca^{2+}$ ions and by quantifying loop opening relative to the domain core. The local hydrogen-bonding and salt-bridge network surrounding positions 310 and 366 was characterized using distance- and angle-based criteria. Solvent accessibility was evaluated with SASA calculations.

Principal component analysis was performed to identify dominant modes of motion involving the $Ca^{2+}$-binding loops and polybasic membrane-interaction face.

The molecular dynamics simulations revealed distinct structural and dynamic consequences for the D310N and D366E variants relative to wild-type *SYT1* C2B. All systems remained globally stable throughout the trajectories, but the variants differed in the magnitude and distribution of conformational shifts. D310N exhibited the largest departure from the starting structure, with backbone RMSD values consistently exceeding those of both wild-type and D366E across replicate runs. Transient excursions toward 3–3.5 Å were observed in D310N, whereas D366E remained closer to wild-type, typically stabilizing near 2–2.5 Å. These deviations in D310N were concentrated around the $Ca^{2+}$-binding loops, whereas the structural core of the β-sandwich remained intact in all simulations.

Local flexibility analyses demonstrated that D310N induced a pronounced increase in root-mean-square fluctuations within residues flanking the primary $Ca^{2+}$-binding loop. The loss of negative charge at position 310 weakened the electrostatic network that normally stabilizes loop geometry and allowed greater solvent penetration into the region. In contrast, D366E preserved the residue's negative charge and generated only modest increases in flexibility that were confined to residues in the immediate vicinity of position 366. Neither variant produced detectable instability in the distal β-strands, indicating that the effects remained largely localized rather than destabilizing the global fold.

Calcium coordination was differentially affected by the two substitutions. D310N displayed frequent and reproducible disruptions in $Ca^{2+}$-coordinating distances, with intermittent loss of canonical contacts and transient formation of non-native hydrogen bonds involving water or nearby residues. These events resulted in alternate loop conformations that were sampled repeatedly across trajectories. D366E maintained a more stable $Ca^{2+}$-coordinating geometry with only subtle adjustments in side-chain orientation, reflecting the preservation of charge but altered reach of the Glu side chain. Overall, D366E trajectories retained $Ca^{2+}$-bound states similar to wild-type, whereas D310N favored misaligned and more solvent-exposed conformations.

Examination of the hydrogen-bonding and salt-bridge networks supported these observations. In wild-type simulations, Asp310 consistently participated in stabilizing interactions that anchored the $Ca^{2+}$-binding loop. Substitution with Asn eliminated these salt bridges and replaced them with fewer, more transient hydrogen bonds that did not provide equivalent stabilization. Increased water accessibility further disrupted local packing and contributed to the heightened loop mobility. Residue 366 showed a different pattern: although D366E altered the geometry of one or two electrostatic interactions, the overall network remained recognizable, and long-lived contacts were still maintained in most frames.

Principal component analysis demonstrated that the dominant motions of the $Ca^{2+}$-binding loops differed substantially between the variants. Wild-type and D366E occupied overlapping conformational subspaces characterized by coordinated, compact loop movements. By contrast, D310N populated a broader distribution of states enriched in "open" loop conformations in which the $Ca^{2+}$-binding pocket tilted away from the β-sandwich core and presented an expanded solvent-facing surface. These conformations were sampled frequently in D310N and rarely in D366E or wild-type simulations.

Together, these findings indicate that D310N produces a markedly greater perturbation of C2B dynamics than D366E. The combination of increased backbone deviation, elevated loop flexibility, compromised $Ca^{2+}$ coordination, and altered conformational sampling supports a model in which D310N more significantly disrupts the $Ca^{2+}$-dependent structural transitions required for efficient synaptic vesicle fusion and recycling. The milder effects observed for D366E are consistent with partially preserved electrostatics and more limited influence on loop geometry. The gradient of structural disruption observed in the simulations parallels clinical severity and phenotypes observed in experimental models, suggesting that $Ca^{2+}$-binding loop destabilization is a principal driver of variant-specific functional impairment.

**Fig. S1.**

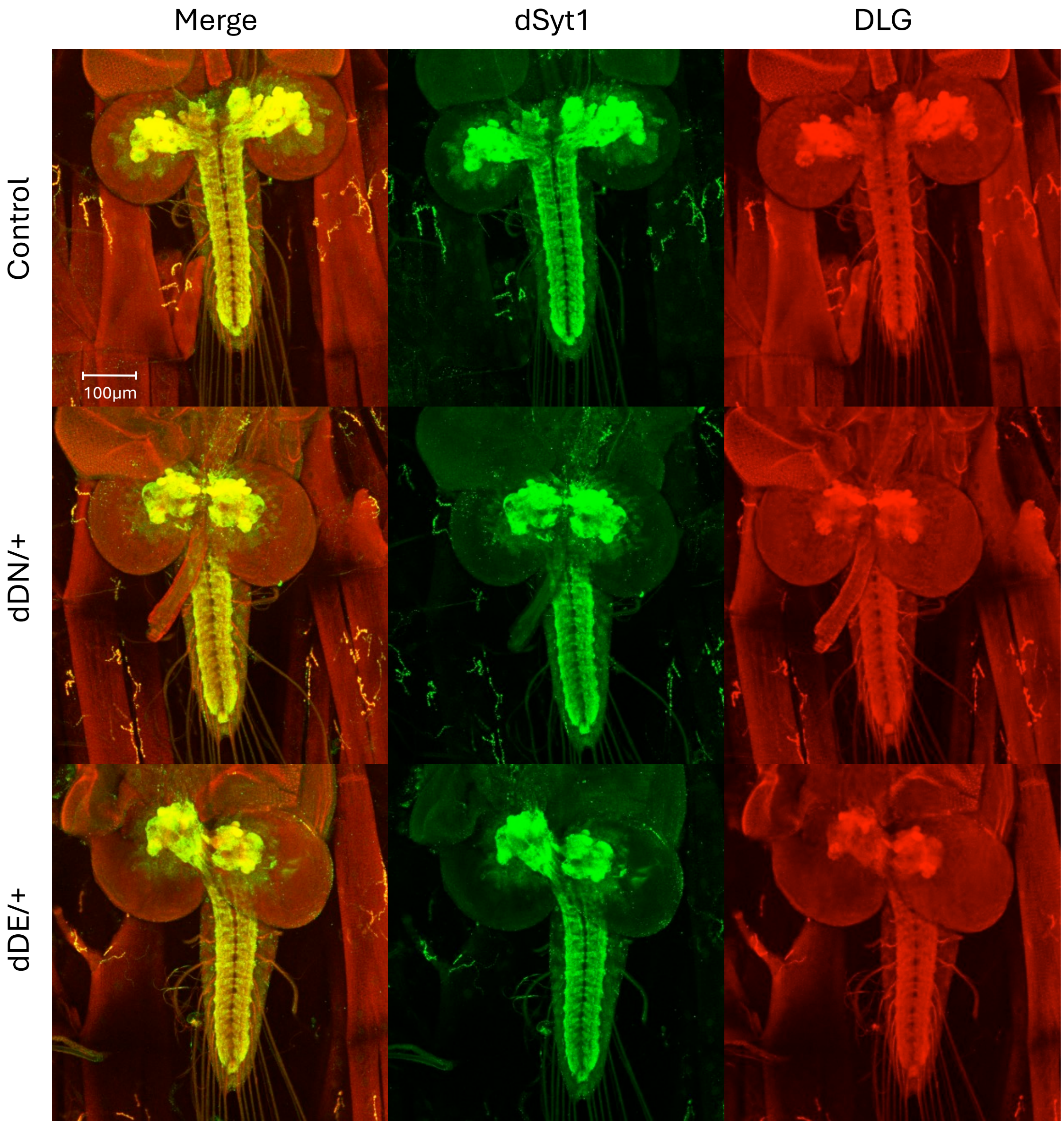


**Supplemental Figure 1. Expression of dSyt1 in heterozygous *dsyt1* mutant larval CNS**

**(A)** The top row shows the representative image of larval brains and ventral verve cords in control animals, stained for dSyt (green) and DLG (red). Note the expression of dSyt1 in neuropils and neuromuscular junctions (NMJs).

**(B)** The middle row shows the representative image of brains and ventral nerve cords in heterozygous *dDN* /+ larvae.

**(C)** The bottom row shows the representative image of brains and ventral nerve cords in heterozygous *dDE* /+ larvae.

Fig. S2.

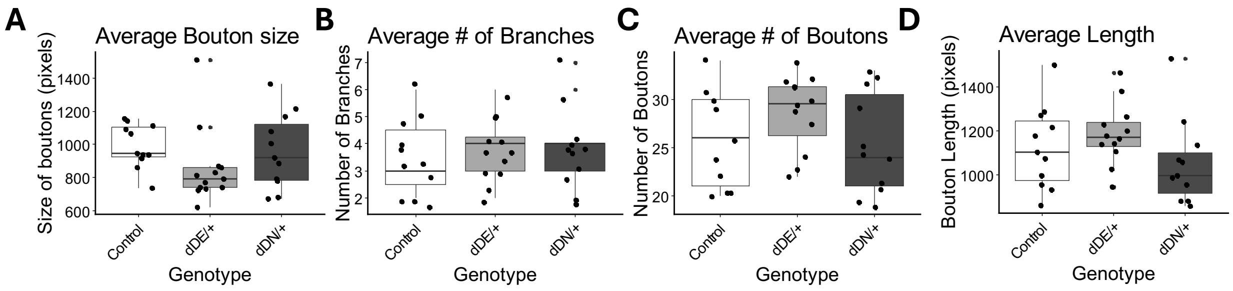


**Supplemental Figure 2. Statistics for NMJ morphology related to Figure 2J**

**(A)** Shows the average bouton size in pixels
**(B)** Shows the average number of branches
**(C)** Shows the average number of boutons
**(D)** Shows the average length across the NMJ

Note there are no statistical differences between any of the parameters measured.

**Fig. S3.**

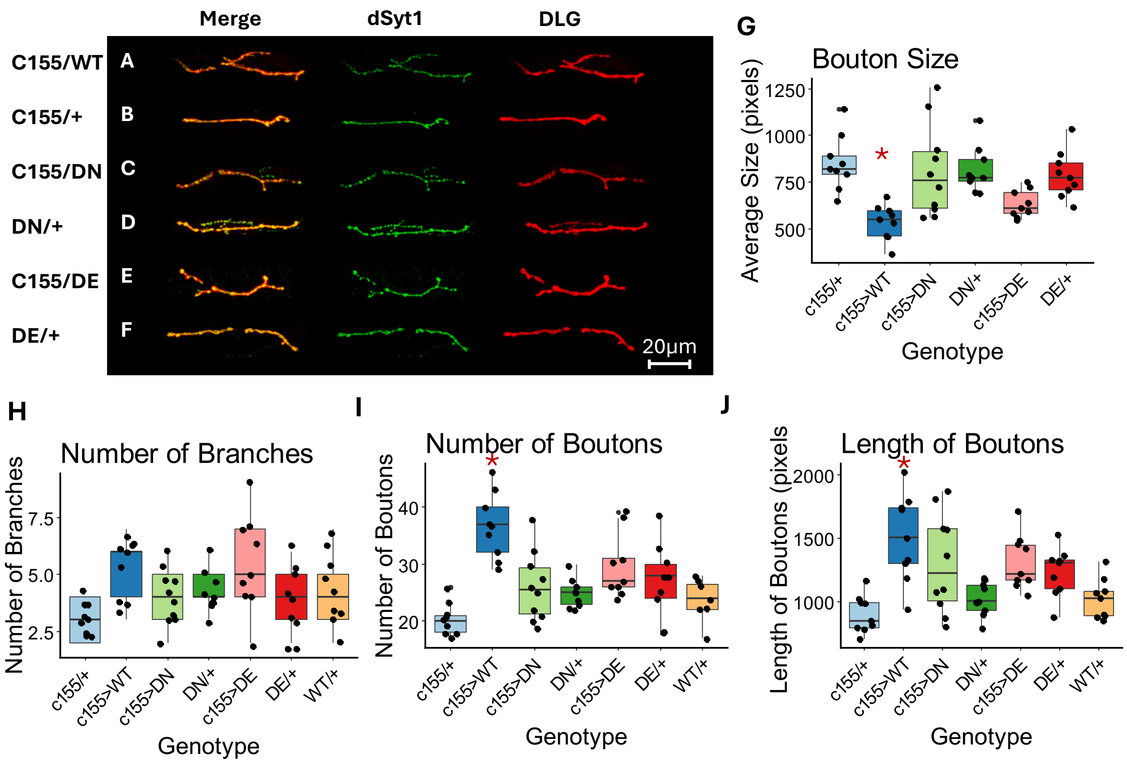


**Supplemental Figure 3. Pan neuronal expression of humanized SYT1 variants does not affect dSYT1 localization and NMJ morphology**

First column shows the merging of the two channels. The second column shows the staining pattern of the native *Drosophila* Syt1 protein (green) at larval NMJs. The last column shows the staining pattern of DLG (red).
C155-Gal4/+, UAS-D366E/+, UAS-D310N/+ serve as controls whereas C155-Gal4>UAS-D310N, C155-Gal4>UAS-D366E and C155-Gal4>UAS-SYT1 WT are experimental groups. Scale bar shown in bottom right corner.

- **(A)** Row shows the C155>UAS-SYT-WT
- **(B)** Row two shows C155-Gal4/+
- **(C)** Row shows C155-Gal4>DN
- **(D)** Row shows UAS-DN/+
- **(E)** Row shows C155-Gal4>DE
- **(F)** Row shows UAS-DE/+
- **(G)** Shows the average bouton size statistics
- **(H)** Shows the number of branches statistics
- **(I)** Shows the number of bouton statistics
- **(J)** Shows the length of bouton statistics

**Fig. S4.**

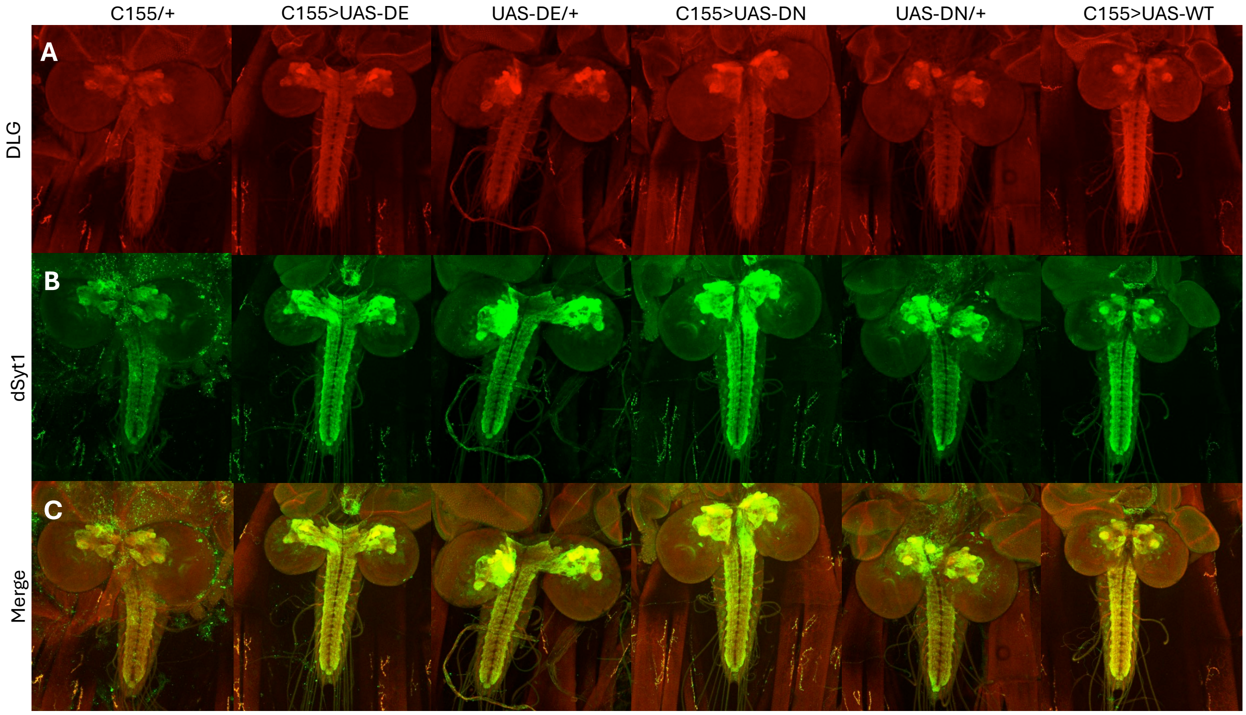


**Supplemental Figure 4. Neuronal expression of humanized mutant *SYT1* does not affect dSyt1 localization in larval CNS**

**(A)** The top row shows the staining pattern of DLG in larval brains and ventral nerve cords (red).

**(B)** The middle row shows the staining pattern of the native *Drosophila* Syt1 (dSyt1) protein in larval brains and ventral nerve cords (green).

**(C)** The bottom row shows the merging of the two channels.

C155-Gal4/+, UAS-D366E/+, UAS-D310N/+ serve as controls whereas C155-Gal4>UAS-D310N, C155-Gal4>UAS-D366E and C155-Gal4>UAS-SYT1 WT are experimental groups.

**Fig. S5.**

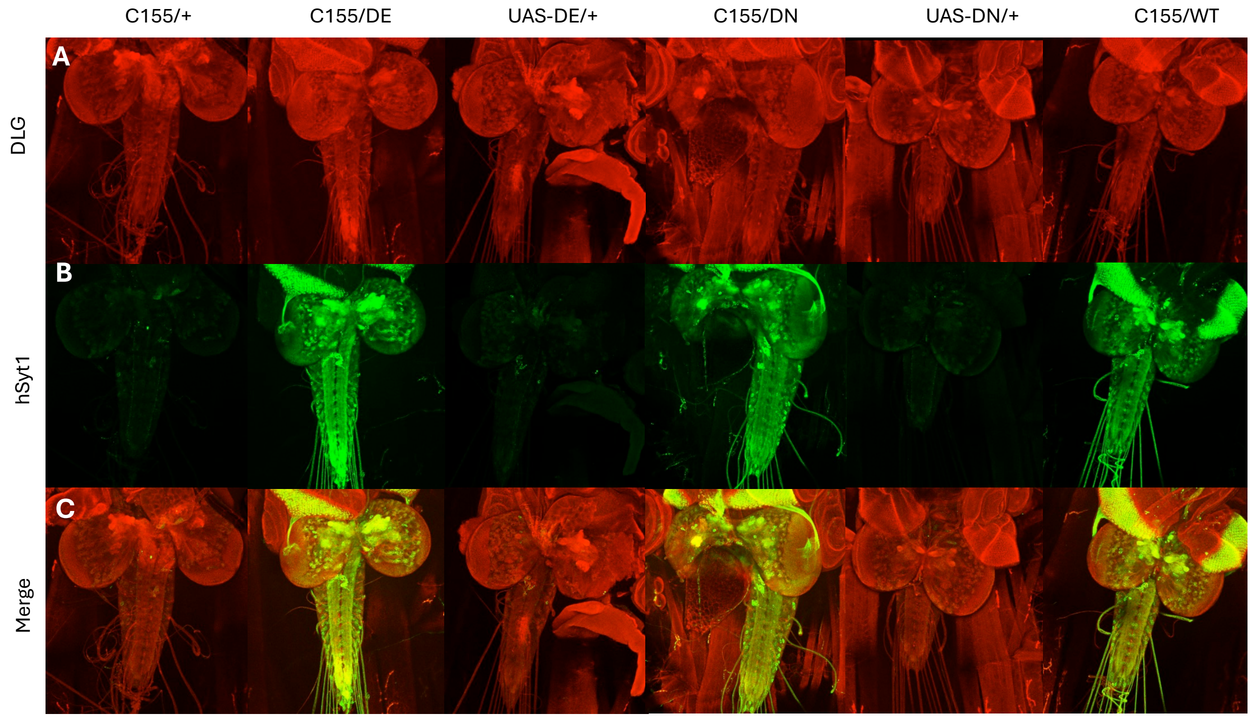


**Supplemental Figure 5. Neuronal expression of humanized mutant *SYT1* does not affect human SYT1 localization in larval CNS**

**(A)** The first row shows the staining pattern of DLG in larval brains and ventral nerve cords (red).

**(B)** The middle row shows the staining pattern of the human SYT1 in larval brains and ventral nerve cords (green).

**(C)** The bottom row shows the merging of the two channels.

C155-Gal4/+, UAS-D366E/+, UAS-D310N/+ serve as controls whereas C155-Gal4>UAS-D310N, C155-Gal4>UAS-D366E and C155-Gal4>UAS-SYT1 WT are experimental groups.

### Table S1.

**Supplemental Table 1. Standardized score ranges and interpretive descriptors used across developmental, adaptive, behavioral, and attentional measures.**

| Standard Score | Scaled Score | T-Score | z-Score | Score Label |
|---|---|---|---|---|
| ≥ 130 | ≥ 16 | ≥ 70 | ≥ + 2.0 | Exceptionally high |
| 120-129 | 14-15 | 64-69 | + 1.4 to 1.9 | Above average |
| 110-119 | 12-13 | 57-63 | + 0.7 to 1.3 | High average |
| 90-109 | 8-11 | 44-56 | ± 0.6 | Average |
| 80-89 | 6-7 | 37-43 | -0.7 to –1.3 | Low average |
| 70-79 | 4-5 | 30-36 | -1.4 to –2.0 | Below average |
| ≤ 69 | ≤ 3 | ≤ 29 | ≤ –2.1 | Exceptionally low |

Standardized score ranges and interpretive descriptors used across developmental, adaptive, behavioral, and attentional measures. Score classifications are derived from age-adjusted normative data provided in the respective test manuals for the Mullen Scales of Early Learning (MSEL), Adaptive Behavior Assessment System, Third Edition (ABAS-3), Child Behavior Checklist (CBCL), Social Responsiveness Scale–Second Edition (SRS-2), and Vanderbilt Assessment Scale.

**Movie S1.**

This video shows the climbing assay and compares *dDE/+* side by side next to control. Compared to the control flies, *dDE/+* flies climb slower and show seizure-like activities after strong taping down (bang) of the graduated cylinder.

**Movie S2.**

This video shows the climbing assay and compares *dDN/+* side by side next to control. Compared to the control flies, *dDN/+* flies climb much slower and show severe seizure-like activities after strong taping down (bang) of the graduated cylinder.

**Movie S3.**

This video shows the control group during the bang-sensitive assay. Following high speed vortexing, the control flies rapidly climb up and show little signs of seizure-like activities.

**Movie S4.**

This video shows the *dDN/+* group during the bang-sensitive assay. Following high speed vortexing, the *dDN/+* flies slowly climb up and show severe seizure-like activities.

**Movie S5.**

This video shows the *dDE/+* group during the bang-sensitive assay. Following high speed vortexing, the *dDE/+* flies climb up but slower than control flies and faster than the *dDN/+* flies. They also show signs of seizure-like activities.

**Movie S6.**

This video shows the $dsyt1^{N6}$/+ (*null/+*) group during the bang-sensitive assay. Following high speed vortexing, the *null/+* flies climb up rapidly and show little or no sign of seizure-like activities.
